# Effectiveness and safety of drugs in pregnancy: evidence from drug target Mendelian randomization

**DOI:** 10.1101/2023.11.06.23298144

**Authors:** Ciarrah-Jane S Barry, Venexia M Walker, Christy Burden, Alexandra Havdahl, Neil M Davies

**Affiliations:** Medical Research Council Integrative Epidemiology Unit, University of Bristol, Bristol, BS82BN, UK; Population Health Sciences, Bristol Medical School, University of Bristol, Bristol, BS82BN, UK; Department of Surgery, University of Pennsylvania Perelman School of Medicine, Philadelphia, Pennsylvania, USA; Translational Health Sciences, Bristol Medical School, University of Bristol, Bristol, UK; Nic Waals Institute, Lovisenberg Diaconal Hospital, Oslo, Norway; Center for Genetic Epidemiology and Mental health, Norwegian Institute of Public Health, Oslo, Norway; PROMENTA, Department of Psychology, University of Oslo, Oslo, Norway; K.G. Jebsen Center for Genetic Epidemiology, Department of Public Health and Nursing, Norwegian University of Science and Technology, Trondheim, Norway; Division of Psychiatry, University College London, London, UK; Department of Statistical Science, University College London, London, UK

**Keywords:** Mendelian randomization, pharmacoepidemiology, genetics, drugs, hypertension, pregnancy

## Abstract

Limited information exists regarding the impact of pharmacotherapy in pregnancy due to ethical concerns of unintended foetal harm. We investigate genetically proxied intrauterine antihypertensive exposure on offspring outcomes, including gestational age and birthweight, using two-sample multivariable Mendelian randomization. Higher levels of maternal protein targets for calcium channel blockers increased gestational age by 3.99 days (95%CI: 0.02, 7.96) per 10mmHg decrease in SBP. Genetically proxied maternal protein targets for beta-adrenoceptor blocking drugs, vasodilator antihypertensive drugs on the KNCJ11 gene, potassium-sparing diuretics and aldosterone antagonists demonstrated little evidence of increased risk to offspring. Both parental genetic protein targets for vasodilator antihypertensive drugs demonstrated similar effects on birthweight, suggesting detrimental offspring effects due to genetic perturbation of these pathways is unlikely. Little evidence for increased risk of adverse offspring outcomes due to maternal antihypertensive drug target perturbation was found. Triangulation of these findings with existing evidence may guide physicians and mothers during pregnancy.

## Background

Pre-existing chronic and acute conditions may require therapeutic management during pregnancy to avoid adverse maternal outcomes. Approximately 8-10% of pregnancies in the UK are affected by hypertension or high blood pressure, including chronic hypertension (pre-existing, typically essential), gestational hypertension (new after 20 weeks gestation), and pre-eclampsia (hypertension with additional features of multiorgan involvement). When left untreated, these are well-established risk factors for numerous serious adverse maternal and early infant outcomes (1,2). These include preeclampsia, maternal death, preterm birth, intrauterine growth restriction, low birth weight and neuropsychiatric disorders (1–3). Yet, treatments for these conditions have been associated with possible additional risks for the developing foetus (**Supplementary Table 1**).

Profound physiologic changes occur during pregnancy, such as increased body weight, renal blood flow and cardiac output (4–6). These have been found and theorised to impact the pharmacokinetics of many drugs, affecting the distribution, absorption, and metabolism (5–7). Yet, clinical trials typically exclude pregnant women. Recruiting pregnant women to randomised controlled trials (RCTs) is challenging for ethical and practical reasons. Additionally, clinical trials using pregnant animals are of limited relevance as there may be species-specific effects, whereby a drug is harmful in some animal species but not in a human or vice-versa (8). As a result, there is relatively little evidence about the effects of drugs in pregnancy, both on women and their offspring. Observational studies on pregnant women have found evidence of possible impaired neonatal development due to drug exposure (teratogenic effects) (9–11). However, the data is limited and often conflicting. Clinical guidance is developed using limited available pharmacological evidence, typically with a tendency towards conservative behaviour. Thus, fear of unintentional foetal harm potentially puts mothers and their offspring at risk through medication avoidance or non-adherence (12). In the UK, the most widely used and recommended antihypertensive drug in pregnancy is labetalol (beta-adrenoreceptor blocker), but its use has been associated with foetal growth restriction and neonatal hypoglycaemia (11,13,14). Nifedipine (calcium channel blocker) is the second most used antihypertensive drug in pregnancy, followed by methyldopa, neither of which is routinely recommended out of pregnancy. Typically, evidence has shown low adherence to antihypertensive medication in pregnancy, which can put the mother and foetus at risk (15,16).

Genetics can provide a complementary source of evidence about the effects of drugs in utero. Mendelian randomization (MR) is an instrumental variables analysis in which genetic variants associated with the exposure of interest are used to assess the causal relationships between an exposure and an outcome (17). A genetic variant fulfils the criteria of a valid instrument if it (a) is reliably associated with the exposure of interest (relevance) (b), has no uncontrolled common cause with the outcome relationship (independence) and (c), only associates with the outcome via its effect on the exposure of interest (exclusion restriction).

An advantage of MR is that it can assess drug safety within pregnancy without exposing the foetus to additional risks. Most drugs work by changing the expression of proteins in the body (18). Drug target MR uses genetic variants within genes that affect proteins targeted by a drug (19). Conventional MR uses genetic variants in unrelated individuals to estimate the effects of manipulating a protein in an individual. However, here, we are interested in the intrauterine effects of drugs taken by the mother during pregnancy. Thus, we want to know the effect of manipulating the protein in the mother on the offspring outcome. We can estimate these effects using intergenerational within-family MR in large datasets of genotyped parent-offspring trios. If the MR assumptions hold, the maternal genotype is a proxy for prescription drug exposure in utero.

Genetic variants are randomly transmitted from parents to offspring at conception and thus cannot be affected by external factors after conception. However, we note exposure-related segregation distortion may exist, meaning the probability of a genetic variant being transmitted from parent to offspring could be influenced by environmental factors (17,20,21). Without related segregation distortion, this random allocation of ‘genetic exposure’ is analogous to the randomisation of treatment within an RCT. As a result, conditional on parental genotype and in the absence of selection bias, offspring genotypes will be independent of the pre-conception environment.

We may instrument drug exposure using genetic variants within genes that relate to the activity of or encode the protein target of the drug (19). Intergenerational within-family MR uses genetic variants in one generation (e.g., the mother’s genotype) as an instrument for maternal exposure to estimate the effects on the offspring (22,23). Thus, maternal genetic variants related to the activity or expression of a drug target or biomarker may be used to proxy drug exposure effects on infant outcomes to determine evidence of potential teratogenic or beneficial effects, Figure 1 (19,24).

**Figure 1:**
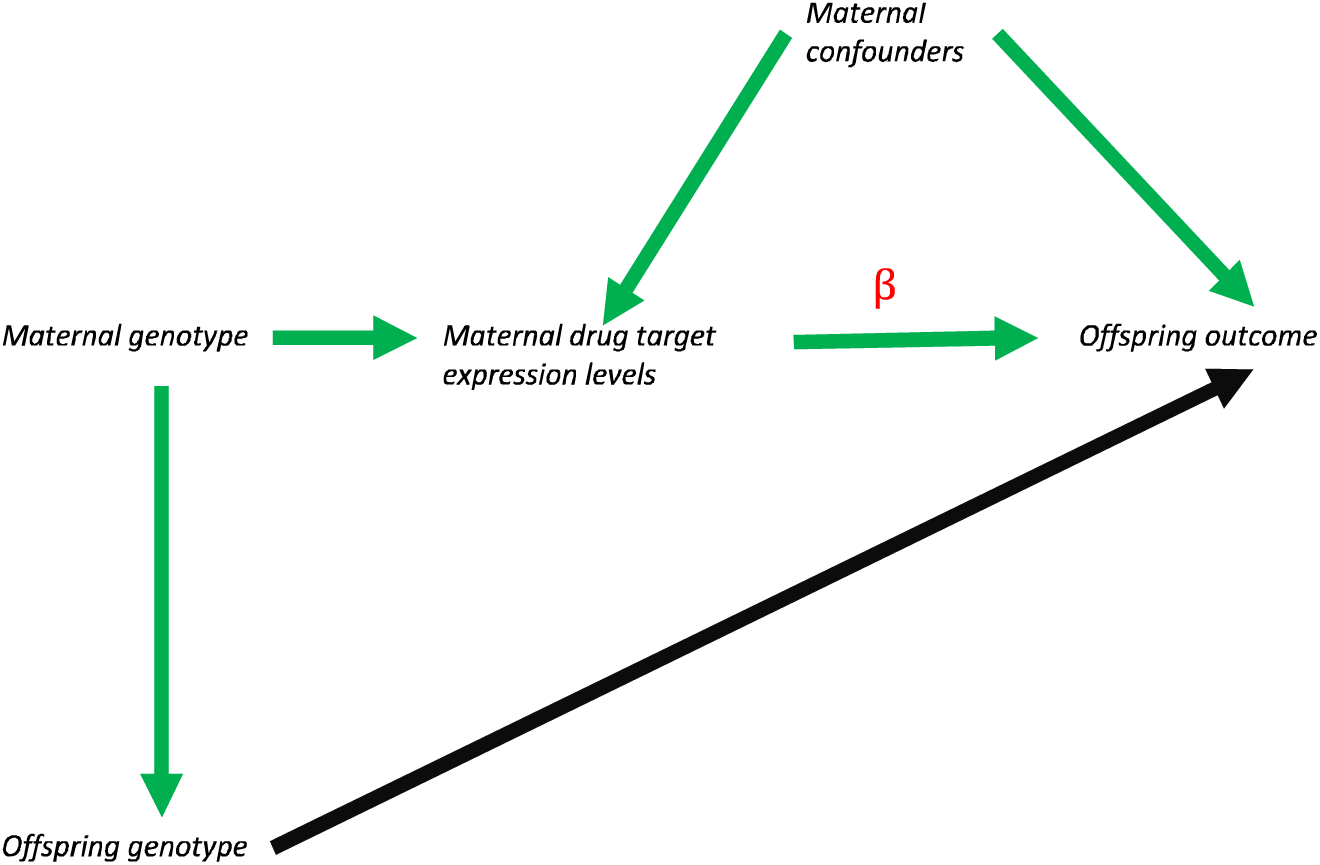
A directed acyclic graph (DAG) demonstrating the intergenerational within family pharmacological MR framework. The black arrows indicate offspring effects mediated via phenotypic expression in the offspring. Green arrows demonstrate the genetic inheritance of the offspring from its mother. β represents the estimate of interest. Here, maternal exposure to a drug target of interest, such as a beta-adrenoreceptor blocker, is instrumented by the maternal genotype in a gene, for example, the single-nucleotide polymorphisms (SNPs) on the ADRB1 gene targeted by the drug substance. The offspring genotype may directly influence the offspring outcome, thus an estimate that controls for offspring genotype will be unbiased by direct genetic inheritance as it would close the direct causal pathway and ensure the independence assumption holds.

MR has been implemented in the literature to identify opportunities for drug repurposing, drug targets and predicting adverse drug effects (19,25–30). However, few studies have used MR to estimate the intrauterine effects of drugs. Evidence from MR could be triangulated with findings from other study designs to guide clinical decision-making, develop or repurpose drugs or establish drug safety profiles.

In this study, we developed and implemented a novel instrument derivation method to determine whether we may predict the causal effects of intrauterine antihypertensive prescriptive drug exposure on offspring outcomes.

## Methods

### Study design

We implemented a two-sample MR analysis to determine the impact of genetically proxied intrauterine drug exposure on the offspring. Summary level SNP-exposure associations were extracted from the IEU Open GWAS platform (31). These estimates were calculated within a sample of 436,419 male and female European participants and were not adjusted for measures of weight or medication use (32,33).

We derived the SNP-outcome associations using individual-level data from the Norwegian Mother, Father and Child Cohort Study (MoBa), a prospective population-based pregnancy cohort study conducted by the Norwegian Institute of Public Health. Pregnant women were recruited at approximately week 18 of gestation across Norway between 1999-2008 (34,35). The women consented to participation in 41% of the invited pregnancies. The cohort includes approximately 114,761 children, 95,248 mothers and 74,626 fathers. The establishment of MoBa and initial data collection was based on a license from the Norwegian Data Protection Agency and approval from The Regional Committees for Medical and Health Research Ethics. The Norwegian Health Registry Act currently regulates the MoBa cohort. The current study was approved by The Regional Committees for Medical and Health Research Ethics (2017/1702).

Data collection occurred at multiple time points within the pregnancy in the form of self-reported questionnaires that were continued after birth. English translations of the questionnaires are available online (36). The MoBa data in this study uses version 12 of the quality-assured data, released in January 2019. Additional information regarding the child’s birth record is available from linkage to the Medical Birth Registry of Norway (MBRN), the national compulsory registry containing information about all births in Norway from 1967 (34,35,37). Blood samples were obtained from both parents during pregnancy and from mothers and children (umbilical cord) at birth (38). Genetic data is available for 44,017 mother-father-child trios, with details on genotyping, imputation and quality control available elsewhere (39).

The specific data of interest to this study were from birth information from the MBRN, questionnaire 6 months after birth and genotyped data from mother, father, and offspring trios. We restricted the sample to parent-offspring trios with complete genetic and outcome covariate data, **Supplementary** Figure 1 and **Supplementary Table 2**.

### Exposures

Using the dm+d search on OpenPrescribing hypertension related virtual medicinal products (VMP) were determined via British National Formulary (BNF) code (40,41). BNF codes and corresponding drug subclasses determined to be relevant to hypertension are listed in **Table 1**.

**Table 1:** BNF codes used to search the NHS dm+d to determine drug substances relevant to the study.

| BNF code | Drug subclass(es) |
| --- | --- |
| 020201 | Thiazides and related diuretics |
| 020202 | Loop diuretics |
| 020203 | Potassium-sparing diuretics and aldosterone antagonists |
| 02040 | Beta-adrenoceptor blocking drugs |
| 02050 | Vasodilator antihypertensive drugs, centrally-acting antihypertensive drugs, adrenergic neurone blocking drugs, alpha-adrenoceptor blocking drugs, renin-angiotensin system drugs, angiotensin-converting enzyme inhibitors, angiotensin-II receptor antagonists, renin inhibitors, ganglion-blocking drugs, other adrenergic neurone blocking drugs |
| 020602 | Calcium channel blockers |
| 040102 | Anxiolytics |

*dm+d; dictionary of medicines and devices, BNF; British National Formulary, NHS; National Health Service,*

Drug substances known to be prescribed for hypertension without allocated BNF code in the OpenPrescribing dm+d were subsequently manually added (meprobamate, potassium chloride, tadalafil, tamsulosin).

### Instrument selection

The active drug substance was identified in each exposure VMP, **Supplementary Table 3**. For each drug substance, the corresponding pharmacologically active gene was identified in DrugBank (42). Using GENCODE, each gene was mapped to its corresponding genome location (chromosome:base pair range) as indicated by release 43 of the GRCh37 assembly (43). All SNPs within these genomic regions were extracted from the MoBa trio-dataset from each member (mother, father, offspring).

### Outcomes

A binary measure for “hypertensive disorders of pregnancy” was derived as a positive control. “Hypertensive disorders of pregnancy” was set to “yes” if there was evidence within the following MBRN variables: hypertension in pregnancy, eclampsia, preeclampsia, early preeclampsia, Haemolysis, Elevated Liver enzymes and Low Platelets syndrome (HELLP) (44).

The Ages and Stages Questionnaire items in the 6-month questionnaire was used to calculate an offspring developmental score, details in **Supplementary note 1**. Thus, the outcomes of interest were hypertensive disorders of pregnancy, and offspring birthweight (g), gestational age (days), birth length (cm), head circumference (cm), Apgar score at 1 minute, Apgar score at 5 minutes and developmental score at 6 months.

Each SNP-outcome association was estimated using linear or logistic regression for continuous or binary measures. To ensure the independence assumption was met, paternal and offspring genotypes were included in the regression alongside the maternal genotype.

### Exposure data

Summary-level data from a genome-wide association study (GWAS) of SBP within UK Biobank was used as the exposure dataset. The sample contained 436,419 male and female participants of European ancestry. We selected the variants that were common to both MoBa and UK Biobank. We then identified the subset of SNPs that were associated with systolic blood pressure 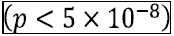, and then clumped with a linkage disequilibrium threshold of r^2^<0.01.

### Statistical methods

We used a two-sample multivariable MR analysis to estimate the effect of the derived maternal drug proxies on early infant outcomes. We used R (version 4.3.0) to analyse the data via the TwoSampleMR package (45,46). The datasets were harmonised, and palindromic SNPs that were non-inferable from their allele frequency were discarded (SNPs having a minor allele frequency > 0.42). Estimates were obtained per 10mmHg lower systolic blood pressure (SBP) as this is comparable to the effect of taking an antihypertensive (47).

We estimated the effects of a 10mmHg lower SBP on each outcome using the Wald estimator for drug classes with a single SNP. For drug subclasses with multiple SNPs, we first checked the mechanism of action of all genes targeted by each SNP for the drug subclass. If the mechanisms of action were identical, we used the inverse variance weighted (IVW) estimator for each outcome. If the mechanisms of action were conflicting, SNPs were subdivided into their mechanistic groupings. Coefficient estimates for binary measures were exponentiated and reported as causal odds ratios (OR).

### Sensitivity analysis

To determine whether the SNPs were acting on the offspring via the maternal genotype, we used the paternal genotype as a negative control (48). We estimated the paternal effect on the offspring outcome whilst controlling for the maternal and offspring genotypes. If the effects of variants on the children’s outcomes was due to the intrauterine environment, then we would expect the maternal, but not the paternal variants to associate with the outcomes.

Additionally, we tested the relevance assumption by calculating the individual and mean F-statistics of the instrument-exposure association. An F-statistic greater than 10 is indicative that the model is unlikely to suffer from substantial weak instrument bias (49).

## Results

### Instrument derivation

The overall instrument derivation process is displayed in **Figure 2**.

**Figure 2:**
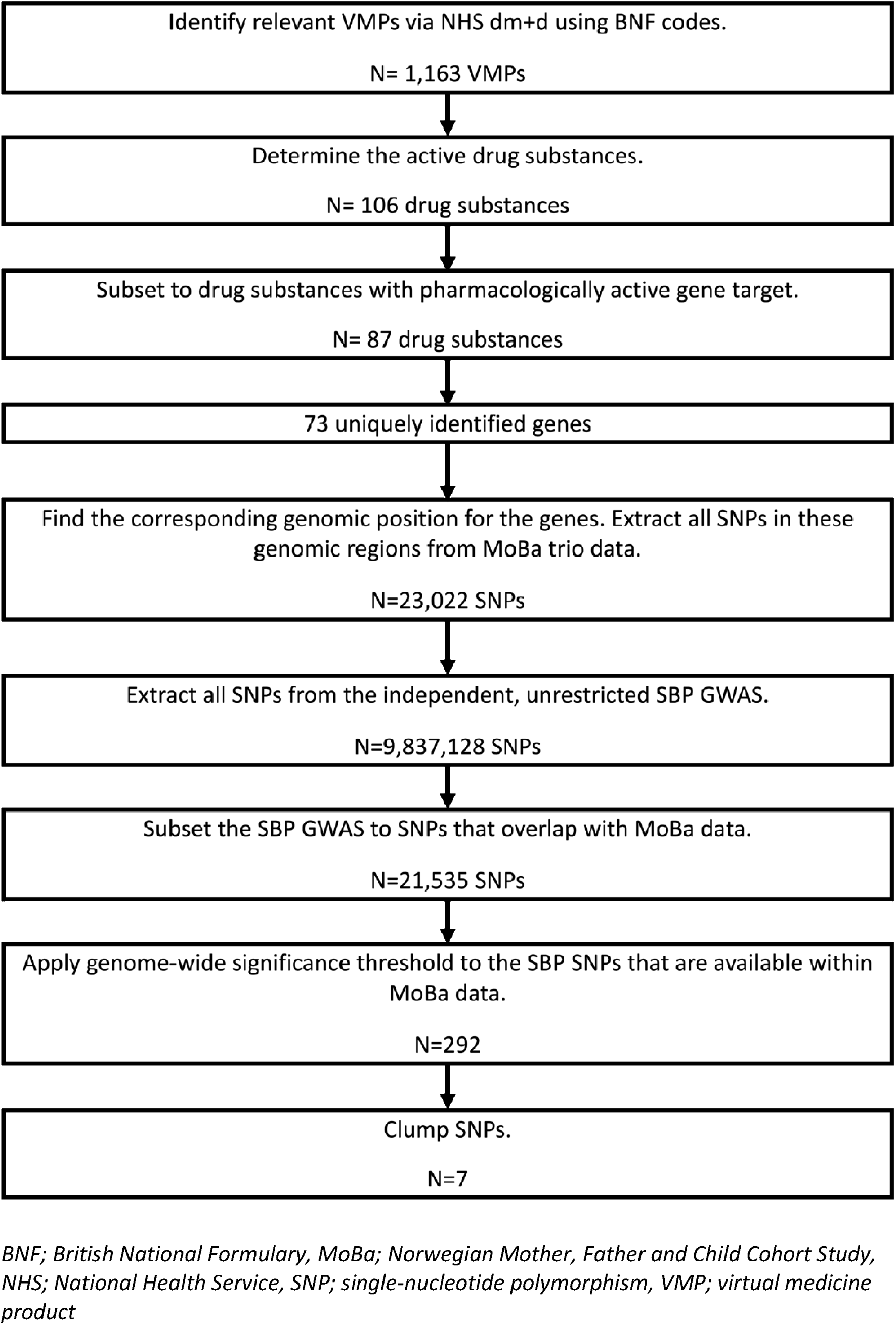
A flowchart describing the instrument derivation procedure.

Within this study, instruments were selected from 23,022 SNPs in 73 uniquely identified genomic regions. The unrestricted SNP-exposure SBP GWAS contained 9,837,128 SNPs. 21,535 SNPs were available in both the SNP-exposure and SNP-outcome datasets. After applying the GWAS p-value threshold, 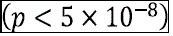, 292 SNPs remained. These SNPs were then clumped to ensure the estimation of statistically independent signals. After all exclusions, we obtained 7 SNPs as instruments.

### Intrauterine effects

Results for the effect of the maternal genotype on offspring outcomes are displayed in **Figure 3**, **Supplementary Table 4**.

**Figures 3:**
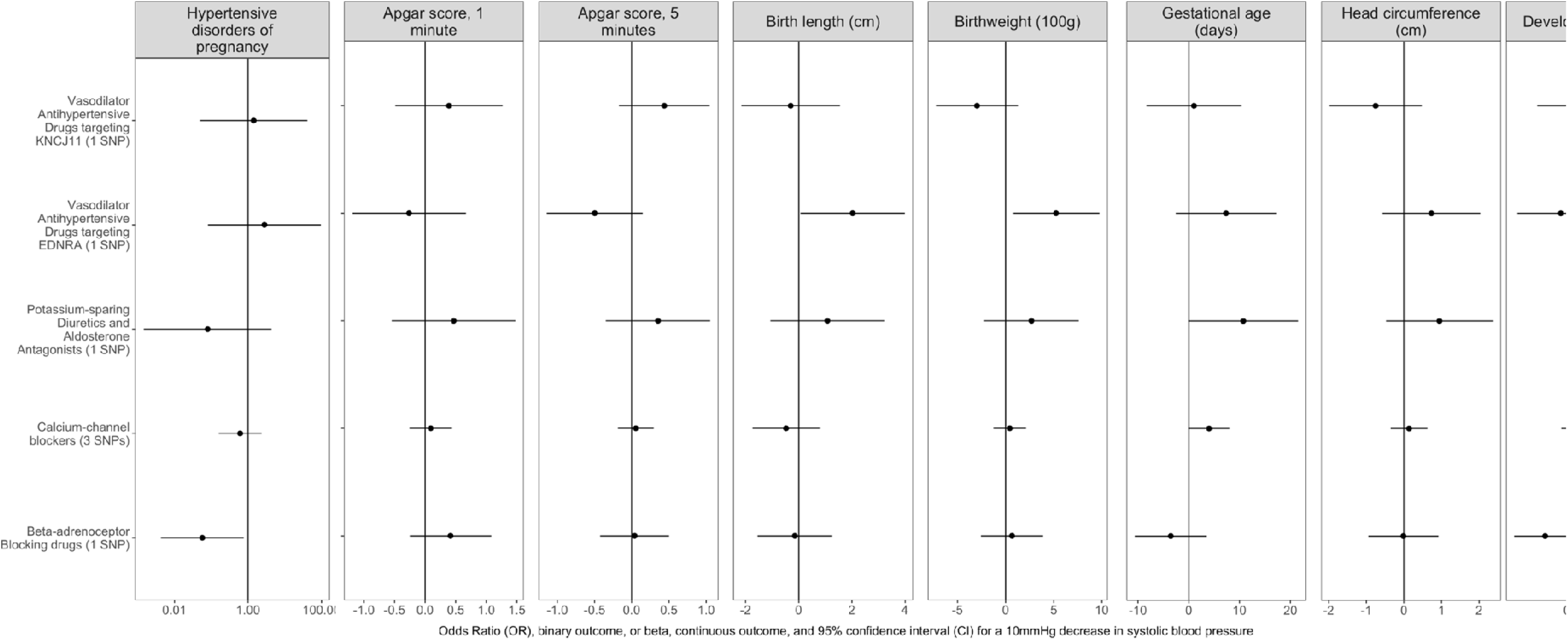
A forest plot demonstrating the estimated causal effect of the maternal genetic drug target on offspring outcomes. Results are shown for the IVW estimate where multiple SNPs were available and the Wald ratio otherwise. An odds ratio (OR) has been estimated for the outcome of “hypertensive disorders of pregnancy”. All other estimates are mean differences.

Across all available drug subclasses, there was some evidence from our analyses of the effects of genetically proxied maternal antihypertensive protein targets on early infant outcomes. Higher levels of vasodilator antihypertensive drugs targeting the EDRNA gene demonstrated evidence of effect on some early infant outcomes. A 10mmHg decrease in SBP resulted in an increase in birthweight of 524g (95% CI: 73, 974); and an increase birth length of 2.03cm (95% CI: 0.06, 3.99). However, we found little evidence that this maternal protein target substantially affected gestational age (7.34 days older (95% CI: –2.55, 17.22) per 10mmHg decrease in SBP). Further, we estimated increased odds of hypertensive disorders of pregnancy, OR 49.29 (1.44, 1686.54), how this estimate was imprecise.

Genetically proxied maternal protein targets for potassium-sparing diuretics and aldosterone antagonists on the SCNN1D gene increased the raw offspring developmental score at 6 months by 0.95 (95% CI: 0.25, 1.65) per 10mmHg decrease in SBP. This estimate was directionally consistent with gestational age (10.73 days, 95% CI: –0.08, 21.53, per 10mmHg decrease in SBP).

Genetically proxied maternal protein targets for vasodilator antihypertensive drugs on the KNCJ11 gene and beta-adrenoceptor blocking drugs suggested little increased risk of adverse early infant outcomes. Higher levels of maternal protein targets for vasodilator antihypertensive drugs on the KNCJ11 gene were estimated to reduce birthweight by 297g per 10mmHg decrease in SBP (95% CI: –129, 723). This suggests there are unlikely to be substantial effects of these maternal protein targets on the early infant outcomes within our study.

We found little evidence that the genetically proxied maternal protein targets for calcium channel blockers affect other early infant outcomes. However, there was evidence that higher levels of these maternal protein targets increased gestational age, with an estimated increase of 3.99 days (95% CI: 0.02, 7.96) per 10mmHg decrease in SBP.

## Sensitivity analysis

### Paternal effect estimates

The effects of paternal protein levels on offspring outcomes are displayed in **Supplementary** Figure 2 and **Supplementary Table 5**. We found little evidence that the genetic paternal protein targets for vasodilator antihypertensive drugs on the KNCJ11 gene and beta-adrenoceptor blocking drugs affected early infant outcomes. However, higher levels of vasodilator antihypertensive drugs targeting EDRNA gene increased birthweight by 485g (95% CI: 32, 937) per 10mmHg decrease in SBP.

Paternal genetic protein targets for potassium-sparing diuretics and aldosterone antagonists decreased birthweight by 647g (95% CI: 147, 1146) per 10mmHg decrease in SBP. Additionally, they reduced head circumference by 1.63cm (95% CI: 0.18, 3.07) per 10mmHg decrease in SBP.

Paternal genetic protein targets for calcium channel blockers increased offspring developmental score at 6 months by 0.28 (0.04, 0.52) per 100mmHg decrease in SBP.

### Offspring effect estimates

The effects of offspring protein levels on offspring outcomes are displayed in **Supplementary Table 6** for completeness.

### Instrument strength

F-statistics were calculated for the individual instruments and, where multiple instruments were available, averaged across the drug class Supplementary Table 7. All F-statistics were greater than 10, and mean F-statistics ranged between 38.39 (vasodilator antihypertensive drugs targeting KCNJ11) and 90.32 (calcium channel blockers targeting the CACNB2 and CACNA1D genes).

## Discussion

In this study, we investigated the effects of drug targets for hypertensive conditions in pregnancy on early infant outcomes. We derived novel summary data for early infant outcomes in MoBa and, with publicly available summary data from the IEU OpenGWAS database for the exposure, applied intergenerational within family MR. We found some evidence that higher levels of maternal drug targets for treatments may improve early infant outcomes. Our results for most outcomes were precise and suggest that many of the drug protein targets, such as those for beta-adrenoreceptor blocking drugs, are unlikely to have large detrimental effects on the infant outcomes within this study. Further, our negative control exposure, paternal levels of drug targets, suggested that vasodilator antihypertensive drugs targeting EDRNA gene had similar effects in mothers and fathers, suggesting that these associations are unlikely to be due to intrauterine effects. There was evidence that some genetically proxied maternal and paternal drugs targets differentially affected offspring outcomes.

No antihypertensive within our positive control analysis demonstrated a reduced risk of maternal hypertensive disorders of pregnancy. However, our estimates were imprecise. Furthermore, these instruments were not selected as antihypertensive drug targets within a pregnant population, and the mechanisms through which they act may differ.

Our results could be explained by horizontal pleiotropy (50), which occurs when a genetic variant influences both the exposure (e.g., intrauterine drug exposure) and the outcome (e.g. neonatal birthweight) through pathways other than the drug target of interest, violating the exclusion restriction criteria. This bias can be toward or away from the null (22,50).

The drug target genetic variants that may influence the offspring outcome via direct inheritance. However, we adjusted for the offspring genetic variants, which should control for direct genetic effects (22).

Assortative mating creates associations for a number of heritable phenotypes between maternal and paternal genotypes (51). Typically, this inflates the estimated effect through violation of the exclusion restriction criteria if there is assortment on the exposure of interest. However, mothers and fathers are unlikely to assort on protein levels or blood pressure (which is normally unobserved in younger populations). Thus, assortative mating is unlikely to substantially impact our analyses. Evidence from analysis of siblings suggests that biological traits (e.g., c-reactive protein) are less likely to be biased by population or familial effects (52).

Population stratification, the systematic difference in allele frequencies and phenotypes between subset of populations, may also confound MR estimates (53). Here the estimated causal genetic to phenotypic effects may be spurious associations explained by ancestral differences (53,54). MoBa is a large, relatively homogenous European sample therefore it is unlikely confounding due to population stratification greatly biases our findings (35,55). Additionally, we adjusted for the top 20 principal components to minimise confounding due to residual population stratification (53,55). Furthermore, population structure is likely to affect maternal and paternal variants equally; thus cannot explain differential parental findings between mothers and fathers.

We investigated the effects of drug targets as proxied by genetic variants known to affect proteins targeted by these drugs. We cannot be certain that these genetic proxies for drug exposure will have a comparable effect to taking the drug of interest. However, the mechanism of action is likely to be the same.

MR has been used to predict maternal-neonatal effects within the literature, however, these typically use summary-level data from unrelated individuals and do not link maternal-exposure-neonatal outcomes or have genetic data from parent-offspring trios. A recent paper implemented MR to investigate the safety of beta-adrenoreceptor-blocking drugs and calcium channel blockers in pregnancy (30). These drug subclasses are typically the most prescribed antihypertensive drugs during pregnancy. Yet there is still little evidence from RCTs to evaluate the risks and benefits of use during pregnancy (56,57). They found evidence to suggest that genetically proxied beta-adrenoreceptor blockers may reduce birth weight. This relationship has been demonstrated in observational studies and is of clinical concern for healthcare professionals and women when using this medication. Further, they found that genetically proxied calcium channel blockers may reduce the risk of preeclampsia and eclampsia.

We were able to address a limitation of their study through the inclusion of the neonatal and paternal genotypes. This approach meant we could exclude pleiotropic effects via the offspring and father. We found little evidence that these variants affected gestational hypertension (i.e., via preeclampsia or eclampsia). Rather, we demonstrate a birthweight lowering effect of genetically proxied beta-adrenoreceptor blockers may occur through the offspring genotype. In contrast to their study, we found some evidence that increased maternal genetically proxied beta-adrenoreceptor-blocking drug targets lowered the odds of gestational hypertension. Further, we found little evidence that maternal genetically proxied calcium channel blockers affected birthweight.

Our study has several strengths. First, MoBa is a large, extensive, and detailed parent-offspring trio dataset. This enabled novel genetic investigation into potential intrauterine effects whilst controlling for genetic confounding and pleiotropy by including offspring and paternal genotype. Genotypic data has been passed through a strict quality control pipeline, and batch effects and principal components may be adjusted for (39). Additionally, many neonatal outcomes of interest to this study originate from a national birth registry and, thus, are precisely measured with little missingness. Second, MR effect estimates are interpreted as the effect of lifetime exposure (58). In the framework of our study, we aim to mimic the effect of offspring exposure for the duration of the pregnancy to emulate potential long-term exposure to maternal treatment. It is unlikely that maternal levels of these proteins would have large biological effects outside of pregnancy. Third, we implemented two-sample MR with non-overlapping samples, which increased statistical power, alongside reducing the likelihood of ‘winners curse’ and weak instrument bias (59,60). Additionally, in two-sample MR, weak instrument bias is towards the null, avoiding false positive findings (17,60). Fourth, our neonatal outcomes of interest are immediate after birth. Thus, it is temporally impossible for post-natal factors to affect these outcomes, avoiding additional confounding bias. Fifth, for the drug subclass in which we retained multiple genetic instrument we performed IVW analysis to increase precision within estimates and provide greater certainty to our results. The use of multiple SNPs also increases the proportion of variance explained in the exposure by the instrument, increasing statistical power (17). Finally, the instruments used in the SNP-exposure relationship were found to be sufficiently strong.

This study has limitations. Genetic variants exert small lifetime effects relative to typical drug exposure (i.e., larger over a specific period). Therefore, we anticipate our estimated effect sizes may not directly equate to clinical results and encourage interpretation based on the potential direction of effects, not the magnitude. Additionally, individual genetic variants each explain a small proportion of the variation. Thus, although the instruments were above the F-statistic threshold, we may have had insufficient power to detect clinically meaningful effect sizes, however, we could discern conservative evidence of effect for some maternal genetically proxied drug subclasses. Drug target MR is not equivalent to estimating the impact of a pharmacological intervention, rather we focus on interpreting the estimated direction of effect, not magnitude. Our results demonstrated many null findings; thus, we found little evidence of increased offspring risk resulting from increased maternal genetic drug proteins. We could not perform standard MR sensitivity analyses, such as weighted median and weighted mode, to assess the exclusion restriction criterion as these require a larger number of SNPs for the exposure (61). This is a common limitation of drug target MR studies, however, this should be offset against the biological proximity of the genetic variants, which reduces the likelihood of pleiotropic effects (62). Moreover, it is possible that maternal genetic variants affect neonatal outcomes via mechanisms other than protein levels. These unintended off-target effects would not be encapsulated in our estimates.

In addition, this study required large-scale linked familial trio data to control for genetic confounding adequately and have sufficient power to detect effects. After exclusions and quality control, our sample size was reduced, which reduced statistical power. Further, we could not investigate many drug subclasses that were initially identified. However, we had sufficient statistical power to detect effects on many outcomes and could exclude clinically meaningful effects for many negative control outcomes. There are currently few large datasets where genetic for trios and phenotypic information is available for replication. While the power of our study is naturally limited, this study provides proof of concept and a framework for this approach to maternal-neonatal analyses, ideally via large-scale international consortia.

MoBa is subject to selection bias as participants were recruited at the start of their pregnancy. Although their characteristics generally reflect the demography of Norway, mothers are older and in better health than the population (34,35,63). This may limit the generalisability of our findings and introduce collider bias. However, selection is unlikely based on genetic variants for protein levels, which are typically unknown to participants.

## Conclusion

Systematic evidence and guidance regarding the use of prescriptive drugs in pregnancy is severely lacking. Here, our results suggest that several antihypertensive drug targets are unlikely to increase the risk of adverse offspring neonatal outcomes substantially. We have provided a framework for future investigations of the effects of intrauterine drug exposure on neonatal outcomes. This provides an additional source of evidence, which may be triangulated with observational analyses and RCTs, where appropriate, to guide clinical decision-making.

## Declarations

### Ethics approval and consent to participate

This was approved by The Regional Committees for Medical and Health Research Ethics (2017/1702).

### Consent for publication

Not applicable.

### Availability of data and materials

Information on how to access the MoBaPsychGen post-imputation QC data is available here: https://www.fhi.no/en/more/research-centres/psychgen/access-to-genetic-data-after-quality-control-by-the-mobapsychgen-pipeline-v/. The summary level data from IEU OpenGWAS are available https://gwas.mrcieu.ac.uk/datasets/ukb-b-20175/.

### Authors contributions

C.S.B, V.W, N.D conceived the study. V.W, N.D, G.D.S, C.B, A.H supervised the project, advised on study design, interpretation of results and revisions. A.H and C.S.B acquired the data. C.S.B drafted the first manuscript, performed all analysis, and generated statistical figures. All authors read, reviewed, revised, and approved the final manuscript.

## Data Availability

Information on how to access the MoBaPsychGen post-imputation QC data is available here: https://www.fhi.no/en/more/research-centres/psychgen/access-to-genetic-data-after-quality-control-by-the-mobapsychgen-pipeline-v/.

## Acknowledgements

The exposure data used for the analyses described in this manuscript were obtained from the dbGaP Accession phs000424.v8.p2 on 15/03/2022. This work used data from the GENCODE project (https://www.gencodegenes.org/) and the IUE OpenGWAS consortium (https://opengwas.org/). The Norwegian Mother, Father and Child Cohort Study is supported by the Norwegian Ministry of Health and Care services and the Ministry of Education and Research (Kunnskapsdepartementet). We are grateful to all the participating families in Norway who take part in this ongoing cohort study. We thank the Norwegian Institute of Public Health (NIPH) for generating high-quality genomic data. The genotype data was provided by the HARVEST collaboration (supported by the Research Council of Norway (RCN) (#229624), the NORMENT Centre (RCN #223273, South-Eastern Norway Regional Health Authority (SENRHA) and Stiftelsen Kristian Gerhard Jebsen) in collaboration with deCODE Genetics, and the Center for Diabetes Research at the University of Bergen (funded by the ERC AdG project SELECTionPREDISPOSED, Stiftelsen Kristian Gerhard Jebsen, Trond Mohn Foundation, the RCN, the Novo Nordisk Foundation, the University of Bergen, and the Western Norway Regional Health Authority). MoBa analyses were performed on the TSD (Tjeneste for Sensitive Data) facilities, owned by the University of Oslo, operated and developed by the TSD service group at the University of Oslo, IT Department (USIT). The computations/simulations/[SIMILAR] were performed on resources provided by Sigma2 – the National Infrastructure for High-Performance Computing and Data Storage in Norway.

We thank George-Davey Smith for his comments and intellectual contributions to this paper.

## References

1. Laine K, Murzakanova G, Sole KB, Pay AD, Heradstveit S, Räisänen S. Prevalence and risk of pre-eclampsia and gestational hypertension in twin pregnancies: a population-based register study. BMJ Open. 2019 Jul 1;9(7):e029908.

2. Bello NA, Zhou H, Cheetham TC, Miller E, Getahun D, Fassett MJ, et al. Prevalence of Hypertension Among Pregnant Women When Using the 2017 American College of Cardiology/American Heart Association Blood Pressure Guidelines and Association With Maternal and Fetal Outcomes. JAMA Netw Open. 2021 Mar 31;4(3):e213808.

3. Vakil P, Henry A, Craig ME, Gow ML. A review of infant growth and psychomotor developmental outcomes after intrauterine exposure to preeclampsia. BMC Pediatr. 2022 Aug 30;22(1):513.

4. Soma-Pillay P, Catherine NP, Tolppanen H, Mebazaa A, Tolppanen H, Mebazaa A. Physiological changes in pregnancy. Cardiovasc J Afr. 2016;27(2):89–94.

5. Zhao Y, Hebert MF, Venkataramanan R. Basic obstetric pharmacology. Semin Perinatol. 2014 Dec;38(8):475–86.

6. Pariente G, Leibson T, Carls A, Adams-Webber T, Ito S, Koren G. Pregnancy-Associated Changes in Pharmacokinetics: A Systematic Review. PLoS Med. 2016 Nov;13(11):e1002160.

7. Soldin O, Mattison D. Sex Differences in Pharmacokinetics and Pharmacodynamics. Clin Pharmacokinet. 2009;48(3):143–57.

8. Vargesson N. Thalidomide-induced teratogenesis: History and mechanisms. Birth Defects Res. 2015 Jun;105(2):140–56.

9. Walfisch A, Al-maawali A, Moretti ME, Nickel C, Koren G. Teratogenicity of angiotensin converting enzyme inhibitors or receptor blockers. J Obstet Gynaecol. 2011 Aug 1;31(6):465–72.

10. Cooper WO, Hernandez-Diaz S, Arbogast PG, Dudley JA, Dyer S, Gideon PS, et al. Major congenital malformations after first-trimester exposure to ACE inhibitors. N Engl J Med. 2006 Jun 8;354(23):2443–51.

11. Tanaka K, Tanaka H, Kamiya C, Katsuragi S, Sawada M, Tsuritani M, et al. Beta-Blockers and Fetal Growth Restriction in Pregnant Women With Cardiovascular Disease. Circ J Off J Jpn Circ Soc. 2016 Sep 23;80(10):2221–6.

12. Shields KE, Lyerly AD. Exclusion of pregnant women from industry-sponsored clinical trials. Obstet Gynecol. 2013 Nov;122(5):1077–81.

13. National Institute for Health and Care Excellence. Hypertension in pregnancy: diagnosis and management. Hypertens Pregnancy.

14. Bateman BT, Patorno E, Desai RJ, Seely EW, Mogun H, Maeda A, et al. Late Pregnancy β Blocker Exposure and Risks of Neonatal Hypoglycemia and Bradycardia. Pediatrics. 2016 Sep;138(3):e20160731.

15. Chen H, Tang Y, Liu C, Liu J, Wang K, Zhang X. Adherence to drug therapy for hypertensive disorders of pregnancy: a cross-sectional survey. Arch Public Health. 2020 May 8;78(1):41.

16. Helou A, Stewart K, George J. Adherence to anti-hypertensive medication in pregnancy. Pregnancy Hypertens. 2021 Aug 1;25:230–4.

17. Burgess S, Davey Smith G, Davies NM, Dudbridge F, Gill D, Glymour MM, et al. Guidelines for performing Mendelian randomization investigations. Wellcome Open Res. 2020 Apr 28;4:186.

18. Bull SC, Doig AJ. Properties of Protein Drug Target Classes. PLoS ONE. 2015 Mar 30;10(3):e0117955.

19. Gill D, Georgakis MK, Walker VM, Schmidt AF, Gkatzionis A, Freitag DF, et al. Mendelian randomization for studying the effects of perturbing drug targets. Wellcome Open Res. 2021;6:16.

20. Sanderson E, Glymour MM, Holmes MV, Kang H, Morrison J, Munafò MR, et al. Mendelian randomization. Nat Rev Methods Primer. 2022 Feb 10;2(1):1–21.

21. Coulton A, Przewieslik-Allen AM, Burridge AJ, Shaw DS, Edwards KJ, Barker GLA. Segregation distortion: Utilizing simulated genotyping data to evaluate statistical methods. PLoS ONE. 2020 Feb 19;15(2):e0228951.

22. Lawlor D, Richmond R, Warrington N, McMahon G, Davey Smith G, Bowden J, et al. Using Mendelian randomization to determine causal effects of maternal pregnancy (intrauterine) exposures on offspring outcomes: Sources of bias and methods for assessing them. Wellcome Open Res. 2017 Feb 14;2:11.

23. Davey Smith G, Ebrahim S. Mendelian randomization: prospects, potentials, and limitations. Int J Epidemiol. 2004 Feb;33(1):30–42.

24. Zheng J, Baird D, Borges MC, Bowden J, Hemani G, Haycock P, et al. Recent Developments in Mendelian Randomization Studies. Curr Epidemiol Rep. 2017;4(4):330– 45.

25. Walker VM, Davey Smith G, Davies NM, Martin RM. Mendelian randomization: a novel approach for the prediction of adverse drug events and drug repurposing opportunities. Int J Epidemiol. 2017 Dec 1;46(6):2078–89.

26. Walker VM, Kehoe PG, Martin RM, Davies NM. Repurposing antihypertensive drugs for the prevention of Alzheimer’s disease: a Mendelian randomization study. Int J Epidemiol. 2020 Aug 1;49(4):1132–40.

27. Khankari NK, Keaton JM, Walker VM, Lee KM, Shuey MM, Clarke SL, et al. Using Mendelian randomisation to identify opportunities for type 2 diabetes prevention by repurposing medications used for lipid management. eBioMedicine [Internet]. 2022 Jun 1 [cited 2022 Sep 16];80. Available from: https://www.thelancet.com/journals/ebiom/article/PIIS2352-3964(22)00219-5/fulltext

28. Bell KJL, Loy C, Cust AE, Teixeira-Pinto A. Mendelian Randomization in Cardiovascular Research. Circ Cardiovasc Qual Outcomes. 2021 Jan;14(1):e005623.

29. Storm CS, Kia DA, Almramhi MM, Bandres-Ciga S, Finan C, Hingorani AD, et al. Finding genetically-supported drug targets for Parkinson’s disease using Mendelian randomization of the druggable genome. Nat Commun. 2021 Dec 20;12(1):7342.

30. Ardissino M, Slob EAW, Rajasundaram S, Reddy RK, Woolf B, Girling J, et al. Safety of beta-blocker and calcium channel blocker antihypertensive drugs in pregnancy: a Mendelian randomization study. BMC Med. 2022 Sep 6;20(1):288.

31. Elsworth B, Lyon M, Alexander T, Liu Y, Matthews P, Hallett J, et al. The MRC IEU OpenGWAS data infrastructure [Internet]. bioRxiv; 2020 [cited 2023 Jun 5]. p. 2020.08.10.244293. Available from: https://www.biorxiv.org/content/10.1101/2020.08.10.244293v1

32. Sudlow C, Gallacher J, Allen N, Beral V, Burton P, Danesh J, et al. UK Biobank: An Open Access Resource for Identifying the Causes of a Wide Range of Complex Diseases of Middle and Old Age. PLOS Med. 2015 Mar 31;12(3):e1001779.

33. Bycroft C, Freeman C, Petkova D, Band G, Elliott LT, Sharp K, et al. The UK Biobank resource with deep phenotyping and genomic data. Nature. 2018 Oct;562(7726):203–9.

34. Magnus P, Birke C, Vejrup K, Haugan A, Alsaker E, Daltveit AK, et al. Cohort Profile Update: The Norwegian Mother and Child Cohort Study (MoBa). Int J Epidemiol. 2016 Apr;45(2):382–8.

35. Magnus P, Irgens LM, Haug K, Nystad W, Skjærven R, Stoltenberg C, et al. Cohort profile: The Norwegian Mother and Child Cohort Study (MoBa). Int J Epidemiol. 2006 Oct 1;35(5):1146–50.

36. Updated P. Norwegian Institute of Public Health. [cited 2022 Feb 23]. Questionnaires from MoBa. Available from: https://www.fhi.no/en/studies/moba/for-forskere-artikler/questionnaires-from-moba/

37. Irgens LM. The Medical Birth Registry of Norway. Epidemiological research and surveillance throughout 30 years. Acta Obstet Gynecol Scand. 2000 Jun;79(6):435–9.

38. Paltiel L, Anita H, Skjerden T, Harbak K, Bækken S, Kristin SN, et al. The biobank of the Norwegian Mother and Child Cohort Study – present status. Nor Epidemiol [Internet]. 2014 Dec 22 [cited 2023 Jul 10];24(1–2). Available from: https://www.ntnu.no/ojs/index.php/norepid/article/view/1755

39. Corfield EC, Frei O, Shadrin AA, Rahman Z, Lin A, Athanasiu L, et al. The Norwegian Mother, Father, and Child cohort study (MoBa) genotyping data resource: MoBaPsychGen pipeline v.1 [Internet]. bioRxiv; 2022 [cited 2023 Jul 10]. p. 2022.06.23.496289. Available from: https://www.biorxiv.org/content/10.1101/2022.06.23.496289v3

40. NHSBSA. NHS Dictionary of Medicines and Devices (dm+d) Data Model R2 v3.2 February 2023 [Internet]. NHSBSA; 2023 [cited 2023 Jun 6]. Available from: https://www.nhsbsa.nhs.uk/pharmacies-gp-practices-and-appliance-contractors/dictionary-medicines-and-devices-dmd

41. About | OpenPrescribing [Internet]. [cited 2022 Sep 23]. Available from: https://openprescribing.net/about/

42. Wishart DS, Feunang YD, Guo AC, Lo EJ, Marcu A, Grant JR, et al. DrugBank 5.0: a major update to the DrugBank database for 2018. Nucleic Acids Res. 2018 Jan 4;46(Database issue):D1074–82.

43. Frankish A, Diekhans M, Ferreira AM, Johnson R, Jungreis I, Loveland J, et al. GENCODE reference annotation for the human and mouse genomes. Nucleic Acids Res. 2019 Jan 8;47(Database issue):D766–73.

44. Al-Khatib A, Sagot P, Cottenet J, Aroun M, Quantin C, Desplanches T. Major postpartum haemorrhage after frozen embryo transfer: A population-based study. BJOG Int J Obstet Gynaecol [Internet]. [cited 2023 Sep 4];n/a(n/a). Available from: https://onlinelibrary.wiley.com/doi/abs/10.1111/1471-0528.17625

45. Hemani G, Zheng J, Elsworth B, Wade KH, Haberland V, Baird D, et al. The MR-Base platform supports systematic causal inference across the human phenome. eLife. 7:e34408.

46. R Core Team. R: A Language and Environment for Statistical Computing. Vienna;

47. Law MR, Wald NJ, Morris JK, Jordan RE. Value of low dose combination treatment with blood pressure lowering drugs: analysis of 354 randomised trials. BMJ. 2003 Jun 26;326(7404):1427.

48. Davey Smith G, Lipsitch M, Tchetgen ET, Cohen T. Negative Control Exposures in Epidemiologic Studies. Epidemiology. 2012;23(2):350–2.

49. Staiger D, Stock JH. Instrumental Variables Regression with Weak Instruments. Econometrica. 1997;65(3):557–86.

50. Hemani G, Bowden J, Davey Smith G. Evaluating the potential role of pleiotropy in Mendelian randomization studies. Hum Mol Genet. 2018 Aug 1;27(R2):R195–208.

51. Hartwig FP, Davies NM, Davey Smith G. Bias in Mendelian randomization due to assortative mating. Genet Epidemiol. 2018 Oct;42(7):608–20.

52. Howe LJ, Nivard MG, Morris TT, Hansen AF, Rasheed H, Cho Y, et al. Within-sibship genome-wide association analyses decrease bias in estimates of direct genetic effects. Nat Genet. 2022 May;54(5):581–92.

53. Brumpton B, Sanderson E, Heilbron K, Hartwig FP, Harrison S, Vie GÅ, et al. Avoiding dynastic, assortative mating, and population stratification biases in Mendelian randomization through within-family analyses. Nat Commun. 2020 Jul 14;11(1):3519.

54. Price AL, Zaitlen NA, Reich D, Patterson N. New approaches to population stratification in genome-wide association studies. Nat Rev Genet. 2010 Jul;11(7):459–63.

55. Hernáez Á, Rogne T, Skåra KH, Håberg SE, Page CM, Fraser A, et al. Body mass index and subfertility: multivariable regression and Mendelian randomization analyses in the Norwegian Mother, Father and Child Cohort Study. Hum Reprod Oxf Engl. 2021 Dec;36(12):3141.

56. Bone JN, Sandhu A, Abalos ED, Khalil A, Singer J, Prasad S, et al. Oral Antihypertensives for Nonsevere Pregnancy Hypertension: Systematic Review, Network Meta– and Trial Sequential Analyses. Hypertens Dallas Tex 1979. 2022 Mar;79(3):614–28.

57. Abalos E, Duley L, Steyn DW, Gialdini C. Antihypertensive drug therapy for mild to moderate hypertension during pregnancy. Cochrane Database Syst Rev. 2018 Oct 1;2018(10):CD002252.

58. Sanderson E, Richardson TG, Morris TT, Tilling K, Davey Smith G. Estimation of causal effects of a time-varying exposure at multiple time points through multivariable mendelian randomization. PLoS Genet. 2022 Jul;18(7):e1010290.

59. Burgess S, Davies NM, Thompson SG. Bias due to participant overlap in two-sample Mendelian randomization. Genet Epidemiol. 2016 Nov;40(7):597–608.

60. Lawlor DA. Commentary: Two-sample Mendelian randomization: opportunities and challenges. Int J Epidemiol. 2016 Jun;45(3):908–15.

61. Skrivankova VW, Richmond RC, Woolf BAR, Davies NM, Swanson SA, VanderWeele TJ, et al. Strengthening the reporting of observational studies in epidemiology using mendelian randomisation (STROBE-MR): explanation and elaboration. BMJ. 2021 Oct 26;375:n2233.

62. Holmes MV, Richardson TG, Ference BA, Davies NM, Davey Smith G. Integrating genomics with biomarkers and therapeutic targets to invigorate cardiovascular drug development. Nat Rev Cardiol. 2021 Jun;18(6):435–53.

63. Tapia G, Størdal K, Mårild K, Kahrs CR, Skrivarhaug T, Njølstad PR, et al. Antibiotics, acetaminophen and infections during prenatal and early life in relation to type 1 diabetes. Int J Epidemiol. 2018 Oct 1;47(5):1538–48.

