## Supplementary materials for "Effectiveness and safety of drugs in pregnancy: evidence from drug target Mendelian randomization"

### Supplementary tables

Supplementary table 1: A non-exhaustive review of the literature describing adverse neonatal outcomes following exposure to the indicated drugs/drug subclasses. Note, this includes a range of retrospective, case-series, case-studies and observational studies that may be underpowered due to a lack of RCTs and other systematic evidence.

| **Drug/drug subclass** | **Adverse neonatal outcome** |
| --- | --- |
| Beta-adrenergic receptor antagonists | Impaired foetal growth, neonatal hypoglycaemia (1–4) |
| Alpha adrenergic blocking agents (ARB) | Congenital malformations, fetopathy (5–7) |
| Angiotensin-converting enzyme (ACE) inhibitors | Fetopathy, congenital malformations, miscarriage (5–8) |
| Calcium channel blockers (CCB) | Seizures (3,8) |
| Vasodilators | Hypertrichosis (9) |

Supplementary table 2: Summary statistics for the study outcome measures within the Norwegian Mother, Father and Child Cohort Study (MoBa).

| **Variable** | **Mean (SD) or count (%)** | **NA (N)** |
| --- | --- | --- |
| Hypertensive disorders of pregnancy |  | 0 |
| Yes | 1703 (5.70) |  |
| No | 28196 (94.3) |  |
| Gestational age | 279.32 (12.18) | 128 |
| Head circumference (cm) | 35.28 (1.61) | 543 |
| Apgar score at 1 minute | 8.69 (1.13) | 59 |
| Apgars score at 5 minutes | 9.43 (0.79) | 58 |
| Birth length (cm) | 50.36 (2.4) | 1150 |
| Birthweight (100g) | 35.95 (55.88) | 44 |
| Developmental score | 9.45 (0.73) | 4400 |

SD; standard deviation

Supplementary table 3: A table of the drug substances, corresponding genes and BNF codes used in the instrument selection procedure.

| **Drug substance** | **DrugBank ID** | **Gene** | **BNF Code** |
| --- | --- | --- | --- |
| Acebutolol | DB01193 | ADRB1 | 204000 |
| Aliskiren | DB09026 | REN | 205053 |
| Ambrisentan | DB06403 | EDNRA | 205010 |
| Amiloride | DB00594 | SCNN1A | 202030 |
| Amiloride | DB00594 | SCNN1D | 202030 |
| Amiloride | DB00594 | SCNN1G | 202030 |
| Amiloride | DB00594 | SCNN1B | 202030 |
| Amlodipine | DB00381 | CACNA1C | 206020 |
| Amlodipine | DB00381 | CACNA1I | 206020 |
| Atenolol | DB00335 | ADRB1 | 204000 |
| Azilsartan Medoxomil | DB08822 | AGTR1 | 205052 |
| Bendroflumethiazide | DB00436 | SLC12A3 | 202010 |
| Bendroflumethiazide | DB00436 | KCNMA1 | 202010 |
| Betaxolol | DB00195 | ADRB1 | 204000 |
| Bisoprolol | DB00612 | ADRB1 | 204000 |
| Bosentan | DB00559 | EDNRA | 205010 |
| Bosentan | DB00559 | EDNRB | 205010 |
| Bumetanide | DB00887 | SLC12A4 | 202020 |
| Bumetanide | DB00887 | SLC12A1 | 202020 |
| Bumetanide | DB00887 | SLC12A2 | 202020 |
| Bumetanide | DB00887 | SLC12A5 | 202020 |
| Candesartan | DB13919 | AGTR1 | 205052 |
| Captopril | DB01197 | ACE | 205051 |
| Carvedilol | DB01136 | ADRB1 | 20400080 |
| Carvedilol | DB01136 | ADRA1A | 20400080 |
| Carvedilol | DB01136 | ADRA1B | 20400080 |
| Carvedilol | DB01136 | ADRA1D | 20400080 |
| Celiprolol | DB04846 | ADRB2 | 20400060 |
| Celiprolol | DB04846 | ADRB1 | 20400060 |
| Chlorothiazide | DB00880 | CA1 | 202010 |
| Chlorothiazide | DB00880 | CA2 | 202010 |
| Chlorothiazide | DB00880 | SLC12A3 | 202010 |
| Chlortalidone | DB00310 | CA1 | 202010 |
| Chlortalidone | DB00310 | SLC12A1 | 202010 |
| Cilazapril | DB01340 | ACE | 205051 |
| Clonidine | DB00575 | ADRA2B | 205020 |
| Clonidine | DB00575 | ADRA2C | 205020 |
| Clonidine | DB00575 | ADRA2A | 205020 |
| Debrisoquine | DB04840 | SLC6A2 | 205030 |
| Diazoxide | DB01119 | KCNJ11 | 205010 |
| Diazoxide | DB01119 | CA2 | 205010 |
| Diazoxide | DB01119 | CA1 | 205010 |
| Diltiazem | DB00343 | CACNG1 | 206020 |
| Diltiazem | DB00343 | CACNA1C | 206020 |
| Doxazosin | DB00590 | ADRA1D | 205040 |
| Doxazosin | DB00590 | ADRA1A | 205040 |
| Enalapril | DB00584 | ACE | 205051 |
| Eplerenone | DB00700 | NR3C2 | 202030 |
| Eprosartan | DB00876 | AGTR1 | 205052 |
| Felodipine | DB01023 | CACNA1S | 206020 |
| Felodipine | DB01023 | CACNB2 | 206020 |
| Felodipine | DB01023 | CACNA2D1 | 206020 |
| Felodipine | DB01023 | CACNA1D | 206020 |
| Felodipine | DB01023 | CACNA1C | 206020 |
| Finerenone | DB16165 | NR3C2 | 202030 |
| Fosinopril | DB00492 | ACE | 205051 |
| Furosemide | DB00695 | SLC12A1 | 202020 |
| Guanethidine | DB01170 | SLC6A2 | 205030 |
| Guanfacine | DB01018 | ADRA2A | 205020 |
| Hydralazine | DB01275 | AOC3 | 205010 |
| Hydrochlorothiazide | DB00999 | KCNMA1 | 202010 |
| Hydrochlorothiazide | DB00999 | SLC12A3 | 202010 |
| Iloprost | DB01088 | PTGIR | 205010 |
| Iloprost | DB01088 | PTGER1 | 205010 |
| Indapamide | DB00808 | SLC12A3 | 202010 |
| Irbesartan | DB01029 | AGTR1 | 205052 |
| Isradipine | DB00270 | CACNA1H | 206020 |
| Isradipine | DB00270 | CACNA2D1 | 206020 |
| Isradipine | DB00270 | CACNA1D | 206020 |
| Isradipine | DB00270 | CACNB2 | 206020 |
| Isradipine | DB00270 | CACNA1S | 206020 |
| Isradipine | DB00270 | CACNA2D2 | 206020 |
| Isradipine | DB00270 | CACNA1C | 206020 |
| Labetalol | DB00598 | ADRB1 | 204000 |
| Labetalol | DB00598 | ADRB2 | 204000 |
| Labetalol | DB00598 | ADRA1B | 204000 |
| Labetalol | DB00598 | ADRA1A | 204000 |
| Labetalol | DB00598 | ADRA1D | 204000 |
| Lacidipine | DB09236 | CACNB1 | 206020 |
| Lacidipine | DB09236 | CACNB2 | 206020 |
| Lacidipine | DB09236 | CACNB3 | 206020 |
| Lacidipine | DB09236 | CACNA1S | 206020 |
| Lacidipine | DB09236 | CACNA1A | 206020 |
| Lacidipine | DB09236 | CACNA1D | 206020 |
| Lacidipine | DB09236 | CACNA1F | 206020 |
| Lacidipine | DB09236 | CACNA1C | 206020 |
| Lacidipine | DB09236 | CACNB4 | 206020 |
| Lercanidipine | DB00528 | CACNG1 | 206020 |
| Lisinopril | DB00722 | ACE | 205051 |
| Losartan | DB00678 | AGTR1 | 205052 |
| Macitentan | DB08932 | EDNRA | 205010 |
| Meprobamate | DB00371 | GABRG1 | 401020 |
| Meprobamate | DB00371 | GABRA5 | 401020 |
| Meprobamate | DB00371 | GABRB2 | 401020 |
| Meprobamate | DB00371 | GABRG3 | 401020 |
| Meprobamate | DB00371 | GABRA3 | 401020 |
| Meprobamate | DB00371 | GABRA2 | 401020 |
| Meprobamate | DB00371 | GABRA4 | 401020 |
| Meprobamate | DB00371 | GABRG2 | 401020 |
| Meprobamate | DB00371 | GABRA1 | 401020 |
| Meprobamate | DB00371 | GABRQ | 401020 |
| Meprobamate | DB00371 | GABRB1 | 401020 |
| Meprobamate | DB00371 | GABRE | 401020 |
| Meprobamate | DB00371 | GABRP | 401020 |
| Meprobamate | DB00371 | GABRA6 | 401020 |
| Meprobamate | DB00371 | GABRB3 | 401020 |
| Meprobamate | DB00371 | GABRD | 401020 |
| Methyldopa | DB00968 | DDC | 205020 |
| Methyldopa | DB00968 | ADRA2A | 205020 |
| Metirosine | DB00765 | TH |  |
| Metolazone | DB00524 | SLC12A3 | 202010 |
| Metoprolol | DB00264 | ADRB1 | 204000 |
| Minoxidil | DB00350 | KCNJ1 | 205010 |
| Moexipril | DB00691 | ACE | 205051 |
| Moxonidine | DB09242 | NISCH | 205020 |
| Moxonidine | DB09242 | ADRA2A | 205020 |
| Nadolol | DB01203 | ADRB1 | 204000 |
| Nebivolol | DB04861 | ADRB1 | 204000 |
| Nicardipine | DB00622 | CACNA1C | 206020 |
| Nicardipine | DB00622 | CACNB2 | 206020 |
| Nicardipine | DB00622 | CACNA1D | 206020 |
| Nicardipine | DB00622 | CACNA2D1 | 206020 |
| Nifedipine | DB01115 | CACNA1C | 206020 |
| Nifedipine | DB01115 | CACNB2 | 206020 |
| Nifedipine | DB01115 | CACNA1D | 206020 |
| Nimodipine | DB00393 | CACNA1F | 206020 |
| Nimodipine | DB00393 | CACNA1C | 206020 |
| Nimodipine | DB00393 | CACNB1 | 206020 |
| Nimodipine | DB00393 | CACNB3 | 206020 |
| Nimodipine | DB00393 | CACNB2 | 206020 |
| Nimodipine | DB00393 | CACNA1S | 206020 |
| Nimodipine | DB00393 | CACNA1D | 206020 |
| Nimodipine | DB00393 | CACNB4 | 206020 |
| Nisoldipine | DB00401 | CACNB2 | 206020 |
| Nisoldipine | DB00401 | CACNA1S | 206020 |
| Nisoldipine | DB00401 | CACNA2D1 | 206020 |
| Nisoldipine | DB00401 | CACNA1D | 206020 |
| Nisoldipine | DB00401 | CACNA1C | 206020 |
| Oxprenolol | DB01580 | ADRB1 | 204000 |
| Perhexiline | DB01074 | CPT2 |  |
| Perhexiline | DB01074 | CPT1A |  |
| Phenoxybenzamine | DB00925 | ADRA1A | 205040 |
| Phenoxybenzamine | DB00925 | ADRA2A | 205040 |
| Phentolamine | DB00692 | ADRA1A | 205040 |
| Phentolamine | DB00692 | ADRA2A | 205040 |
| Pindolol | DB00960 | ADRB1 | 204000 |
| Pindolol | DB00960 | ADRB2 | 204000 |
| Polythiazide | DB01324 | SLC12A3 | 202010 |
| Prazosin | DB00457 | ADRA1A | 205040 |
| Prazosin | DB00457 | ADRA1B | 205040 |
| Prazosin | DB00457 | ADRA1D | 205040 |
| Propranolol | DB00571 | ADRB1 | 204000 |
| Quinapril | DB00881 | ACE | 205051 |
| Ramipril | DB00178 | ACE | 205051 |
| Riociguat | DB08931 | GUCY1A2 | 205010 |
| Sildenafil | DB00203 | PDE5A | 205010 |
| Sitaxentan | DB06268 | EDNRA | 205010 |
| Sotalol | DB00489 | KCNH2 | 204000 |
| Sotalol | DB00489 | ADRB2 | 204000 |
| Sotalol | DB00489 | ADRB1 | 204000 |
| Spironolactone | DB00421 | NR3C2 | 202030 |
| Tadalafil | DB00820 | PDE5A | 704050 |
| Tamsulosin | DB00706 | ADRA1A | 704010 |
| Telmisartan | DB00966 | AGTR1 | 205052 |
| Telmisartan | DB00966 | PPARG | 205052 |
| Terazosin | DB01162 | ADRA1B | 205040 |
| Terazosin | DB01162 | ADRA1A | 205040 |
| Terazosin | DB01162 | ADRA1D | 205040 |
| Terazosin | DB01162 | TGFB1 | 205040 |
| Timolol | DB00373 | ADRB1 | 204000 |
| Timolol | DB00373 | ADRB2 | 204000 |
| Torasemide | DB00214 | SLC12A1 | 202020 |
| Torasemide | DB00214 | SLC12A2 | 202020 |
| Trandolapril | DB00519 | ACE | 205051 |
| Triamterene | DB00384 | SCNN1G | 202030 |
| Triamterene | DB00384 | SCNN1B | 202030 |
| Triamterene | DB00384 | SCNN1A | 202030 |
| Valsartan | DB00177 | AGTR1 | 205052 |
| Verapamil | DB00661 | CACNA1C | 206020 |
| Vericiguat | DB15456 | GUCY1B1 | 205010 |
| Olmesartan | DB00275 | AGTR1 | 205052 |
| Perindopril | DB00790 | ACE | 205051 |

Supplementary Table 4: The MR results for the effect of the maternal genetic drug protein targets on offspring outcomes.

| **Outcome** | **Beta coefficient (95% CI)** | **Standard error** | **Pvalue** | **No. SNPs** | **Drug subclass** | **MR method** | **rsID** |
| --- | --- | --- | --- | --- | --- | --- | --- |
| Gestational age (days) | 3.99 (0.02, 7.96) | 3.92 | 0.05 | 3 | Calcium-channel blockers (3 SNPs) | IVW | rs10764331 rs12258967 rs3821843 |
| Birthweight (100g) | 0.43 (-1.26, 2.12) | 167.28 | 0.62 | 3 | Calcium-channel blockers (3 SNPs) | IVW | rs10764331 rs12258967 rs3821843 |
| Birth length (cm) | -0.48 (-1.74, 0.78) | 1.24 | 0.46 | 3 | Calcium-channel blockers (3 SNPs) | IVW | rs10764331 rs12258967 rs3821843 |
| Head circumference (cm) | 0.14 (-0.35, 0.63) | 0.48 | 0.58 | 3 | Calcium-channel blockers (3 SNPs) | IVW | rs10764331 rs12258967 rs3821843 |
| Apgar score, 1 minute | 0.09 (-0.26, 0.44) | 0.34 | 0.6 | 3 | Calcium-channel blockers (3 SNPs) | IVW | rs10764331 rs12258967 rs3821843 |
| Apgar score, 5 minutes | 0.05 (-0.19, 0.29) | 0.24 | 0.66 | 3 | Calcium-channel blockers (3 SNPs) | IVW | rs10764331 rs12258967 rs3821843 |
| Hypertensive disorders of pregnancy | 0.61 (0.16, 2.34) | 1.33 | 0.47 | 3 | Calcium-channel blockers (3 SNPs) | IVW | rs10764331 rs12258967 rs3821843 |
| Developmental score | 0.16 (-0.08, 0.4) | 0.24 | 0.2 | 3 | Calcium-channel blockers (3 SNPs) | IVW | rs10764331 rs12258967 rs3821843 |
| Gestational age (days) | 10.73 (-0.08, 21.53) | 10.68 | 0.05 | 1 | Potassium-sparing Diuretics and Aldosterone Antagonists (1 SNP) | Wald ratio | rs1262894 |
| Gestational age (days) | 7.34 (-2.55, 17.22) | 9.77 | 0.15 | 1 | Vasodilator Antihypertensive Drugs targeting EDNRA (1 SNP) | Wald ratio | rs13143677 |
| Gestational age (days) | 1.03 (-8.32, 10.37) | 9.24 | 0.83 | 1 | Vasodilator Antihypertensive Drugs targeting KNCJ11 (1 SNP) | Wald ratio | rs1557765 |
| Gestational age (days) | -3.54 (-10.63, 3.55) | 7.01 | 0.33 | 1 | Beta-adrenoceptor Blocking drugs (1 SNP) | Wald ratio | rs1801253 |
| Birthweight (100g) | 2.68 (-2.24, 7.6) | 486.33 | 0.29 | 1 | Potassium-sparing Diuretics and Aldosterone Antagonists (1 SNP) | Wald ratio | rs1262894 |
| Birthweight (100g) | 5.24 (0.73, 9.74) | 445.47 | 0.02 | 1 | Vasodilator Antihypertensive Drugs targeting EDNRA (1 SNP) | Wald ratio | rs13143677 |
| Birthweight (100g) | -2.97 (-7.23, 1.29) | 421.17 | 0.17 | 1 | Vasodilator Antihypertensive Drugs targeting KNCJ11 (1 SNP) | Wald ratio | rs1557765 |
| Birthweight (100g) | 0.66 (-2.57, 3.89) | 319.53 | 0.69 | 1 | Beta-adrenoceptor Blocking drugs (1 SNP) | Wald ratio | rs1801253 |
| Birth length (cm) | 1.07 (-1.07, 3.21) | 2.12 | 0.33 | 1 | Potassium-sparing Diuretics and Aldosterone Antagonists (1 SNP) | Wald ratio | rs1262894 |
| Birth length (cm) | 2.03 (0.06, 3.99) | 1.94 | 0.04 | 1 | Vasodilator Antihypertensive Drugs targeting EDNRA (1 SNP) | Wald ratio | rs13143677 |
| Birth length (cm) | -0.3 (-2.16, 1.55) | 1.83 | 0.75 | 1 | Vasodilator Antihypertensive Drugs targeting KNCJ11 (1 SNP) | Wald ratio | rs1557765 |
| Birth length (cm) | -0.15 (-1.55, 1.26) | 1.39 | 0.84 | 1 | Beta-adrenoceptor Blocking drugs (1 SNP) | Wald ratio | rs1801253 |
| Head circumference (cm) | 0.95 (-0.47, 2.36) | 1.4 | 0.19 | 1 | Potassium-sparing Diuretics and Aldosterone Antagonists (1 SNP) | Wald ratio | rs1262894 |
| Head circumference (cm) | 0.73 (-0.57, 2.03) | 1.29 | 0.27 | 1 | Vasodilator Antihypertensive Drugs targeting EDNRA (1 SNP) | Wald ratio | rs13143677 |
| Head circumference (cm) | -0.75 (-1.98, 0.48) | 1.22 | 0.23 | 1 | Vasodilator Antihypertensive Drugs targeting KNCJ11 (1 SNP) | Wald ratio | rs1557765 |
| Head circumference (cm) | -0.01 (-0.95, 0.92) | 0.92 | 0.98 | 1 | Beta-adrenoceptor Blocking drugs (1 SNP) | Wald ratio | rs1801253 |
| Apgar score, 1 minute | 0.47 (-0.55, 1.48) | 1 | 0.37 | 1 | Potassium-sparing Diuretics and Aldosterone Antagonists (1 SNP) | Wald ratio | rs1262894 |
| Apgar score, 1 minute | -0.26 (-1.19, 0.67) | 0.92 | 0.58 | 1 | Vasodilator Antihypertensive Drugs targeting EDNRA (1 SNP) | Wald ratio | rs13143677 |
| Apgar score, 1 minute | 0.39 (-0.49, 1.27) | 0.87 | 0.38 | 1 | Vasodilator Antihypertensive Drugs targeting KNCJ11 (1 SNP) | Wald ratio | rs1557765 |
| Apgar score, 1 minute | 0.41 (-0.25, 1.08) | 0.66 | 0.22 | 1 | Beta-adrenoceptor Blocking drugs (1 SNP) | Wald ratio | rs1801253 |
| Apgar score, 5 minutes | 0.35 (-0.35, 1.05) | 0.69 | 0.33 | 1 | Potassium-sparing Diuretics and Aldosterone Antagonists (1 SNP) | Wald ratio | rs1262894 |
| Apgar score, 5 minutes | -0.5 (-1.14, 0.15) | 0.64 | 0.13 | 1 | Vasodilator Antihypertensive Drugs targeting EDNRA (1 SNP) | Wald ratio | rs13143677 |
| Apgar score, 5 minutes | 0.44 (-0.17, 1.05) | 0.6 | 0.16 | 1 | Vasodilator Antihypertensive Drugs targeting KNCJ11 (1 SNP) | Wald ratio | rs1557765 |
| Apgar score, 5 minutes | 0.04 (-0.43, 0.5) | 0.46 | 0.87 | 1 | Beta-adrenoceptor Blocking drugs (1 SNP) | Wald ratio | rs1801253 |
| Hypertensive disorders of pregnancy | 0.08 (0, 4.35) | 3.96 | 0.21 | 1 | Potassium-sparing Diuretics and Aldosterone Antagonists (1 SNP) | Wald ratio | rs1262894 |
| Hypertensive disorders of pregnancy | 2.79 (0.08, 96.28) | 3.5 | 0.57 | 1 | Vasodilator Antihypertensive Drugs targeting EDNRA (1 SNP) | Wald ratio | rs13143677 |
| Hypertensive disorders of pregnancy | 1.42 (0.05, 41.15) | 3.33 | 0.84 | 1 | Vasodilator Antihypertensive Drugs targeting KNCJ11 (1 SNP) | Wald ratio | rs1557765 |
| Hypertensive disorders of pregnancy | 0.06 (0, 0.76) | 2.57 | 0.03 | 1 | Beta-adrenoceptor Blocking drugs (1 SNP) | Wald ratio | rs1801253 |
| Developmental score | 0.95 (0.25, 1.65) | 0.69 | 0.01 | 1 | Potassium-sparing Diuretics and Aldosterone Antagonists (1 SNP) | Wald ratio | rs1262894 |
| Developmental score | -0.09 (-0.74, 0.56) | 0.64 | 0.78 | 1 | Vasodilator Antihypertensive Drugs targeting EDNRA (1 SNP) | Wald ratio | rs13143677 |
| Developmental score | 0.17 (-0.44, 0.77) | 0.6 | 0.59 | 1 | Vasodilator Antihypertensive Drugs targeting KNCJ11 (1 SNP) | Wald ratio | rs1557765 |
| Developmental score | -0.32 (-0.78, 0.14) | 0.45 | 0.17 | 1 | Beta-adrenoceptor Blocking drugs (1 SNP) | Wald ratio | rs1801253 |

Supplementary Table 5: The MR results for the effect of the paternal genetic drug protein targets on offspring outcomes.

| **Outcome** | **Beta coefficient (95% CI)** | **Standard error** | **Pvalue** | **No. SNPs** | **Drug subclass** | **MR method** | **rsID** |
| --- | --- | --- | --- | --- | --- | --- | --- |
| Gestational age (days) | -2.2 (-9.37, 4.98) | 7.1 | 0.55 | 1 | Beta-adrenoceptor Blocking drugs (1 SNP) | Wald ratio | rs1801253 |
| Birthweight (100g) | -1.08 (-4.35, 2.19) | 323.43 | 0.52 | 1 | Beta-adrenoceptor Blocking drugs (1 SNP) | Wald ratio | rs1801253 |
| Birth length (cm) | -0.55 (-1.97, 0.87) | 1.41 | 0.45 | 1 | Beta-adrenoceptor Blocking drugs (1 SNP) | Wald ratio | rs1801253 |
| Head circumference (cm) | -0.25 (-1.2, 0.69) | 0.93 | 0.6 | 1 | Beta-adrenoceptor Blocking drugs (1 SNP) | Wald ratio | rs1801253 |
| Apgar score, 1 minute | 0.38 (-0.3, 1.05) | 0.67 | 0.27 | 1 | Beta-adrenoceptor Blocking drugs (1 SNP) | Wald ratio | rs1801253 |
| Apgar score, 5 minutes | 0.15 (-0.32, 0.62) | 0.46 | 0.52 | 1 | Beta-adrenoceptor Blocking drugs (1 SNP) | Wald ratio | rs1801253 |
| Hypertensive disorders of pregnancy | 0.29 (0.02, 3.93) | 2.57 | 0.35 | 1 | Beta-adrenoceptor Blocking drugs (1 SNP) | Wald ratio | rs1801253 |
| Developmental score | -0.05 (-0.52, 0.42) | 0.46 | 0.84 | 1 | Beta-adrenoceptor Blocking drugs (1 SNP) | Wald ratio | rs1801253 |
| Gestational age (days) | -8.04 (-19.01, 2.93) | 10.85 | 0.15 | 1 | Potassium-sparing Diuretics and Aldosterone Antagonists (1 SNP) | Wald ratio | rs1262894 |
| Birthweight (100g) | -6.47 (-11.46, -1.47) | 493.92 | 0.01 | 1 | Potassium-sparing Diuretics and Aldosterone Antagonists (1 SNP) | Wald ratio | rs1262894 |
| Birth length (cm) | -1.73 (-3.9, 0.44) | 2.14 | 0.12 | 1 | Potassium-sparing Diuretics and Aldosterone Antagonists (1 SNP) | Wald ratio | rs1262894 |
| Head circumference (cm) | -1.63 (-3.07, -0.18) | 1.43 | 0.03 | 1 | Potassium-sparing Diuretics and Aldosterone Antagonists (1 SNP) | Wald ratio | rs1262894 |
| Apgar score, 1 minute | -0.73 (-1.76, 0.29) | 1.02 | 0.16 | 1 | Potassium-sparing Diuretics and Aldosterone Antagonists (1 SNP) | Wald ratio | rs1262894 |
| Apgar score, 5 minutes | -0.49 (-1.2, 0.23) | 0.7 | 0.18 | 1 | Potassium-sparing Diuretics and Aldosterone Antagonists (1 SNP) | Wald ratio | rs1262894 |
| Hypertensive disorders of pregnancy | 3.3 (0.07, 163.59) | 3.86 | 0.55 | 1 | Potassium-sparing Diuretics and Aldosterone Antagonists (1 SNP) | Wald ratio | rs1262894 |
| Developmental score | 0.11 (-0.6, 0.82) | 0.7 | 0.76 | 1 | Potassium-sparing Diuretics and Aldosterone Antagonists (1 SNP) | Wald ratio | rs1262894 |
| Gestational age (days) | 3.83 (-6.09, 13.76) | 9.81 | 0.45 | 1 | Vasodilator Antihypertensive Drugs targeting EDNRA (1 SNP) | Wald ratio | rs13143677 |
| Birthweight (100g) | 4.85 (0.32, 9.37) | 447.14 | 0.04 | 1 | Vasodilator Antihypertensive Drugs targeting EDNRA (1 SNP) | Wald ratio | rs13143677 |
| Birth length (cm) | 0.55 (-1.42, 2.51) | 1.94 | 0.59 | 1 | Vasodilator Antihypertensive Drugs targeting EDNRA (1 SNP) | Wald ratio | rs13143677 |
| Head circumference (cm) | 0.79 (-0.51, 2.1) | 1.29 | 0.23 | 1 | Vasodilator Antihypertensive Drugs targeting EDNRA (1 SNP) | Wald ratio | rs13143677 |
| Apgar score, 1 minute | -0.22 (-1.16, 0.71) | 0.92 | 0.64 | 1 | Vasodilator Antihypertensive Drugs targeting EDNRA (1 SNP) | Wald ratio | rs13143677 |
| Apgar score, 5 minutes | -0.46 (-1.11, 0.19) | 0.64 | 0.16 | 1 | Vasodilator Antihypertensive Drugs targeting EDNRA (1 SNP) | Wald ratio | rs13143677 |
| Hypertensive disorders of pregnancy | 0.87 (0.03, 29.84) | 3.5 | 0.94 | 1 | Vasodilator Antihypertensive Drugs targeting EDNRA (1 SNP) | Wald ratio | rs13143677 |
| Developmental score | -0.51 (-1.16, 0.14) | 0.64 | 0.12 | 1 | Vasodilator Antihypertensive Drugs targeting EDNRA (1 SNP) | Wald ratio | rs13143677 |
| Gestational age (days) | -3.75 (-13.14, 5.64) | 9.29 | 0.43 | 1 | Vasodilator Antihypertensive Drugs targeting KNCJ11 (1 SNP) | Wald ratio | rs1557765 |
| Birthweight (100g) | -1.53 (-5.81, 2.75) | 423.38 | 0.48 | 1 | Vasodilator Antihypertensive Drugs targeting KNCJ11 (1 SNP) | Wald ratio | rs1557765 |
| Birth length (cm) | 0.4 (-1.46, 2.27) | 1.84 | 0.67 | 1 | Vasodilator Antihypertensive Drugs targeting KNCJ11 (1 SNP) | Wald ratio | rs1557765 |
| Head circumference (cm) | -1.04 (-2.28, 0.19) | 1.22 | 0.1 | 1 | Vasodilator Antihypertensive Drugs targeting KNCJ11 (1 SNP) | Wald ratio | rs1557765 |
| Apgar score, 1 minute | 0.34 (-0.54, 1.22) | 0.87 | 0.45 | 1 | Vasodilator Antihypertensive Drugs targeting KNCJ11 (1 SNP) | Wald ratio | rs1557765 |
| Apgar score, 5 minutes | 0.1 (-0.51, 0.71) | 0.61 | 0.75 | 1 | Vasodilator Antihypertensive Drugs targeting KNCJ11 (1 SNP) | Wald ratio | rs1557765 |
| Hypertensive disorders of pregnancy | 3.13 (0.11, 92.76) | 3.35 | 0.51 | 1 | Vasodilator Antihypertensive Drugs targeting KNCJ11 (1 SNP) | Wald ratio | rs1557765 |
| Developmental score | -0.21 (-0.82, 0.4) | 0.61 | 0.5 | 1 | Vasodilator Antihypertensive Drugs targeting KNCJ11 (1 SNP) | Wald ratio | rs1557765 |
| Gestational age (days) | 3.16 (-3.03, 9.35) | 6.12 | 0.32 | 3 | Calcium-channel blockers (3 SNPs) | IVW | rs10764331 rs12258967 rs3821843 |
| Birthweight (100g) | 0.27 (-1.43, 1.97) | 167.97 | 0.75 | 3 | Calcium-channel blockers (3 SNPs) | IVW | rs10764331 rs12258967 rs3821843 |
| Birth length (cm) | -0.13 (-0.86, 0.61) | 0.73 | 0.74 | 3 | Calcium-channel blockers (3 SNPs) | IVW | rs10764331 rs12258967 rs3821843 |
| Head circumference (cm) | 0.04 (-0.45, 0.53) | 0.49 | 0.86 | 3 | Calcium-channel blockers (3 SNPs) | IVW | rs10764331 rs12258967 rs3821843 |
| Apgar score, 1 minute | -0.03 (-0.38, 0.32) | 0.35 | 0.87 | 3 | Calcium-channel blockers (3 SNPs) | IVW | rs10764331 rs12258967 rs3821843 |
| Apgar score, 5 minutes | -0.09 (-0.33, 0.15) | 0.24 | 0.45 | 3 | Calcium-channel blockers (3 SNPs) | IVW | rs10764331 rs12258967 rs3821843 |
| Hypertensive disorders of pregnancy | 1.16 (0.08, 16.4) | 2.62 | 0.91 | 3 | Calcium-channel blockers (3 SNPs) | IVW | rs10764331 rs12258967 rs3821843 |
| Developmental score | 0.28 (0.04, 0.52) | 0.24 | 0.02 | 3 | Calcium-channel blockers (3 SNPs) | IVW | rs10764331 rs12258967 rs3821843 |

Supplementary Table 6: The MR results for the effect of the offspring genetic drug protein targets on offspring outcomes.

| **Outcome** | **Beta coefficient (95% CI)** | **Standard error** | **Pvalue** | **No. SNPs** | **Drug subclass** | **MR method** | **rsID** |
| --- | --- | --- | --- | --- | --- | --- | --- |
| Gestational age (days) | 8.94 (0.68, 17.2) | 8.17 | 0.03 | 1 | Beta-adrenoceptor Blocking drugs (1 SNP) | Wald ratio | rs1801253 |
| Birthweight (100g) | -4.46 (-8.22, -0.69) | 372.16 | 0.02 | 1 | Beta-adrenoceptor Blocking drugs (1 SNP) | Wald ratio | rs1801253 |
| Birth length (cm) | -0.97 (-2.61, 0.66) | 1.62 | 0.24 | 1 | Beta-adrenoceptor Blocking drugs (1 SNP) | Wald ratio | rs1801253 |
| Head circumference (cm) | -0.77 (-1.86, 0.31) | 1.07 | 0.16 | 1 | Beta-adrenoceptor Blocking drugs (1 SNP) | Wald ratio | rs1801253 |
| Apgar score, 1 minute | -0.51 (-1.28, 0.27) | 0.77 | 0.2 | 1 | Beta-adrenoceptor Blocking drugs (1 SNP) | Wald ratio | rs1801253 |
| Apgar score, 5 minutes | -0.33 (-0.87, 0.2) | 0.53 | 0.22 | 1 | Beta-adrenoceptor Blocking drugs (1 SNP) | Wald ratio | rs1801253 |
| Gestational hypertension | 0.77 (0.04, 13.54) | 2.83 | 0.86 | 1 | Beta-adrenoceptor Blocking drugs (1 SNP) | Wald ratio | rs1801253 |
| Gestational age (days) | -1.34 (-13.84, 11.16) | 12.36 | 0.83 | 1 | Potassium-sparing Diuretics and Aldosterone Antagonists (1 SNP) | Wald ratio | rs1262894 |
| Birthweight (100g) | 2.12 (-3.57, 7.81) | 562.94 | 0.47 | 1 | Potassium-sparing Diuretics and Aldosterone Antagonists (1 SNP) | Wald ratio | rs1262894 |
| Birth length (cm) | 0.18 (-2.29, 2.66) | 2.44 | 0.88 | 1 | Potassium-sparing Diuretics and Aldosterone Antagonists (1 SNP) | Wald ratio | rs1262894 |
| Head circumference (cm) | 0.59 (-1.06, 2.23) | 1.62 | 0.48 | 1 | Potassium-sparing Diuretics and Aldosterone Antagonists (1 SNP) | Wald ratio | rs1262894 |
| Apgar score, 1 minute | 0.13 (-1.04, 1.3) | 1.16 | 0.83 | 1 | Potassium-sparing Diuretics and Aldosterone Antagonists (1 SNP) | Wald ratio | rs1262894 |
| Apgar score, 5 minutes | -0.23 (-1.04, 0.59) | 0.8 | 0.59 | 1 | Potassium-sparing Diuretics and Aldosterone Antagonists (1 SNP) | Wald ratio | rs1262894 |
| Gestational hypertension | 1.17 (0.08, 16.46) | 2.61 | 0.91 | 1 | Potassium-sparing Diuretics and Aldosterone Antagonists (1 SNP) | Wald ratio | rs1262894 |
| Gestational age (days) | -6.07 (-17.41, 5.26) | 11.21 | 0.29 | 1 | Vasodilator Antihypertensive Drugs targeting EDNRA (1 SNP) | Wald ratio | rs13143677 |
| Birthweight (100g) | -3.4 (-8.56, 1.77) | 510.94 | 0.2 | 1 | Vasodilator Antihypertensive Drugs targeting EDNRA (1 SNP) | Wald ratio | rs13143677 |
| Birth length (cm) | -0.91 (-3.15, 1.34) | 2.22 | 0.43 | 1 | Vasodilator Antihypertensive Drugs targeting EDNRA (1 SNP) | Wald ratio | rs13143677 |
| Head circumference (cm) | -0.4 (-1.89, 1.09) | 1.47 | 0.6 | 1 | Vasodilator Antihypertensive Drugs targeting EDNRA (1 SNP) | Wald ratio | rs13143677 |
| Apgar score, 1 minute | 0.01 (-1.05, 1.08) | 1.05 | 0.98 | 1 | Vasodilator Antihypertensive Drugs targeting EDNRA (1 SNP) | Wald ratio | rs13143677 |
| Apgar score, 5 minutes | 0.48 (-0.26, 1.22) | 0.73 | 0.2 | 1 | Vasodilator Antihypertensive Drugs targeting EDNRA (1 SNP) | Wald ratio | rs13143677 |
| Gestational hypertension | 3.11 (0.05, 188.08) | 4.06 | 0.59 | 1 | Vasodilator Antihypertensive Drugs targeting EDNRA (1 SNP) | Wald ratio | rs13143677 |
| Gestational age (days) | -1.51 (-12.34, 9.33) | 10.71 | 0.79 | 1 | Vasodilator Antihypertensive Drugs targeting KNCJ11 (1 SNP) | Wald ratio | rs1557765 |
| Birthweight (100g) | 2.48 (-2.46, 7.42) | 488.26 | 0.32 | 1 | Vasodilator Antihypertensive Drugs targeting KNCJ11 (1 SNP) | Wald ratio | rs1557765 |
| Birth length (cm) | 0.29 (-1.86, 2.43) | 2.12 | 0.79 | 1 | Vasodilator Antihypertensive Drugs targeting KNCJ11 (1 SNP) | Wald ratio | rs1557765 |
| Head circumference (cm) | -0.03 (-1.46, 1.39) | 1.41 | 0.96 | 1 | Vasodilator Antihypertensive Drugs targeting KNCJ11 (1 SNP) | Wald ratio | rs1557765 |
| Apgar score, 1 minute | -0.59 (-1.61, 0.43) | 1.01 | 0.26 | 1 | Vasodilator Antihypertensive Drugs targeting KNCJ11 (1 SNP) | Wald ratio | rs1557765 |
| Apgar score, 5 minutes | -0.32 (-1.02, 0.39) | 0.7 | 0.38 | 1 | Vasodilator Antihypertensive Drugs targeting KNCJ11 (1 SNP) | Wald ratio | rs1557765 |
| Gestational hypertension | 3.94 (0.08, 194.63) | 3.86 | 0.49 | 1 | Vasodilator Antihypertensive Drugs targeting KNCJ11 (1 SNP) | Wald ratio | rs1557765 |
| Gestational age (days) | -5.39 (-10.82, 0.04) | 5.37 | 0.05 | 3 | Calcium-channel blockers (3 SNPs) | IVW | rs10764331 rs12258967 rs3821843 |
| Birthweight (100g) | -1.12 (-3.08, 0.84) | 193.66 | 0.26 | 3 | Calcium-channel blockers (3 SNPs) | IVW | rs10764331 rs12258967 rs3821843 |
| Birth length (cm) | -0.13 (-1.33, 1.06) | 1.18 | 0.83 | 3 | Calcium-channel blockers (3 SNPs) | IVW | rs10764331 rs12258967 rs3821843 |
| Head circumference (cm) | -0.07 (-0.63, 0.5) | 0.56 | 0.82 | 3 | Calcium-channel blockers (3 SNPs) | IVW | rs10764331 rs12258967 rs3821843 |
| Apgar score, 1 minute | -0.03 (-0.43, 0.37) | 0.4 | 0.88 | 3 | Calcium-channel blockers (3 SNPs) | IVW | rs10764331 rs12258967 rs3821843 |
| Apgar score, 5 minutes | 0.05 (-0.23, 0.33) | 0.28 | 0.72 | 3 | Calcium-channel blockers (3 SNPs) | IVW | rs10764331 rs12258967 rs3821843 |
| Gestational hypertension | 14.37 (2.98, 69.29) | 1.56 | 0 | 3 | Calcium-channel blockers (3 SNPs) | IVW | rs10764331 rs12258967 rs3821843 |

Supplementary Table 7: The individual and mean F-statistics for the Wald ratio and IVW estimates respectively to demonstrate the instrument strength of the SNP-exposure relationship.

| **Drug subclass** | **F-statistic** | **No. SNPs** |
| --- | --- | --- |
| Beta-adrenoceptor blocking drugs | 69.23 | 1 |
| Calcium-channel blockers | 90.32 | 3 |
| Potassium-sparing diuretics and aldosterone antagonists | 33.06 | 1 |
| Vasodilator antihypertensive drugs targeting EDNRA | 40.01 | 1 |
| Vasodilator antihypertensive drugs targeting KCNJ11 | 38.39 | 1 |

#### Supplementary notes

Derivation of the prorated developmental score

The Ages and Stages Questionnaire (ASQ) items from the MoBa 6-month questionnaire was used to measure infant neurodevelopmental status. The 11 maternal reported items focus on motor (e.g., “Does your child roll over from his/her back onto his/her tummy?") and communication skills (e.g., “When you call your child, does he/she turn towards you one of the first times you say his/her name?”), with the following response options and scores:

- “Yes, often”, scored 10
- “Yes, but seldom”, scored 5
- “No, not yet”, scored 0
- “Don’t know”, scored as missing

We calculated a prorated total developmental score (10).

Quality control of MoBa data

33,199 individuals in the NORMENT samples were genotyped using Illumina HumanOmniExpress-24v1.0, Illumina InfiniumOmniExpress-24v1.2 and the Illumina Global Screening Array MD v.1.0 + 50k custom OmniExpress overlap content array. 26,990 individuals were genotyped in the ROTTERDAM sample using the Illumina Global Screening Array MD v.1.0 array. 5,410 were sampled in the TED samples using the Illumina InfiniumOmniExpress-24v1.2, and 32,538 were sampled in the HARVEST sample using the llumina HumanCoreExome12v1.1 and Illumina HumanCoreExome24v1.0. Further details regarding pre-imputation QC, phasing and imputation are available elsewhere .

Post-imputation quality control was as follows. Individuals with either reported verses genetic sex mismatch, a sex-chromosome aneuploidy or that were unable to be linked to phenotypic data were excluded from the sample (n=508). The dataset was also checked for Mendelian errors through PLINK’s “—mendel” command, meaning the reported parent was incorrect. A threshold of 1% and 5% for the trio and variant error rate respectively was implemented. This resulted in the removal of a further 129 individuals and 1,293 variants. These SNPs on average were less accurately imputed, with mean INFO=0.88 relative to mean INFO=0.97 for all other SNPs. 3,061 individuals were genotyped twice, and 52 individuals were genotyped 3 times to determine the agreement of SNPs within the pairs of duplicated samples. The 2 samples that were indicated as duplicated by MoBa data had low concordance $\hat{\pi}=0.8$. An additional 2,474 pairs of samples had $0.74<\hat{\pi} <0.98$, however are thought to represent the same individual and not reflect sample contamination. Thus, the heterogeneity is likely a result of the inclusion of the same individuals that were genotyped with different chips that impute to differing standards.

140,767 SNPs were excluded for discordance in more than 5% of the duplicated samples.

Finally, one individual from each pair of duplicated was randomly dropped using a seed .

The sample was restricted to individuals of a “European” ancestry using the first 2 principal components of MoBa data.

The MoBa data were merged with the 1000 Genomes reference panel and the 1000 Genomes principal components were projected onto the MoBa data. The principal component values the MoBa samples were compared to each of the populations included in the reference panel. Samples were excluded if their principal component 1 and 2 values were in the range of the non-European samples in the 1000 Genomes reference panel (n=688) (13).

To determine and exclude based on the degree of relatedness the KING was implemented to estimate kinship coefficients. KING was restricted to an independent set of SNPs with MAF>0.10, window=3000kb and LD R^2^>0.9 and age was included as a covariate.

This identified 86,175 pairs of known related individuals and 10,769 unknown related individuals. Parental relationships were subsequently updated with these results and families were reconstructed. KING updated the family ID for 24,022 individuals and parental relationships for 21,361 individuals. Where newly assigned relationships appeared to be errors, e.g., parents less than 15 years older than their children, both parents of the same sex, individuals that identify as monozygotic twins but are linked to different pregnancies, and siblings with different parents or an age gap exceeding 25 years, samples were flagged for exclusion (n=375). Further, parent-offspring pairs in which the mother or father identified by KING differed to the genotyped individual specified in the pedigree were flagged for exclusion, reflecting sample mix-up or samples in which the sampled partner is not the biological father. Within the samples of parents and offspring GCTA was used to select an unrelated subsample, with identity by state < 5% after pruning to a set of independent HAPMAP3 SNPs (n=14,092 offspring and n=24,836 unrelated parents) (13).

The first 20 principal components were calculated independently on a subset of the data for both parent and offspring, restricting to variants pruned to independence using PLINK in HAPMAP3. Two sets of principal components were constructed, in which the first used the MoBa data accounting for structure within the data and the second was generated using the 1000 genomes reference panel (13).

### Supplementary figures

Supplementary Figure 1: An exclusion flowchart demonstrating the cohort derivation.

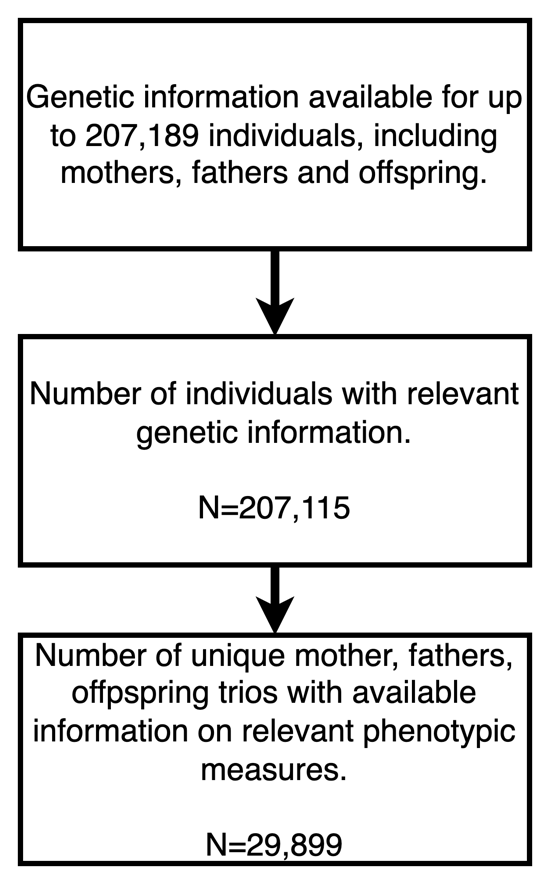

Supplementary Figure 2: A forest plot demonstrating the estimated causal effect of the paternal genetic drug targets on offspring outcomes. Results are shown for the IVW estimate where multiple SNPs were available and Wald ratio otherwise. An odds-ratio (OR) has been estimated for the outcome of “gestational hypertension”. All other estimates are mean differences.

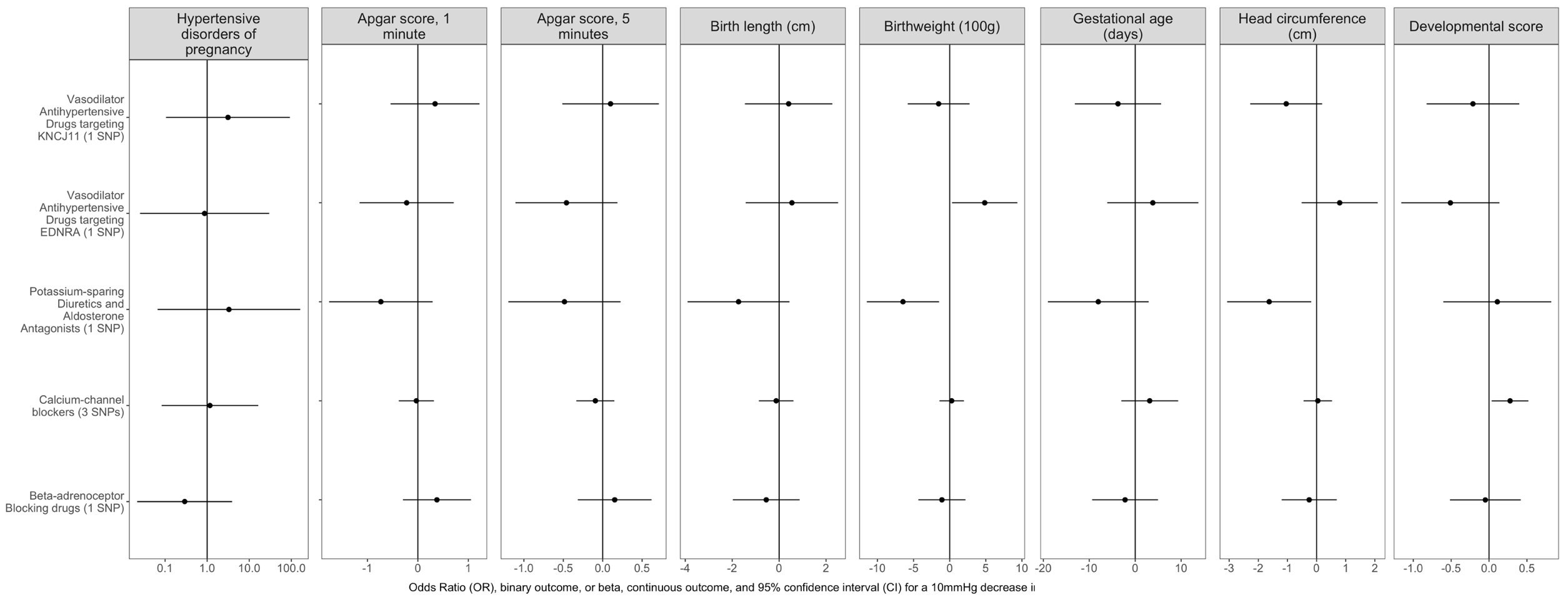

References

1.	Duan L, Ng A, Chen W, Spencer HT, Lee MS. Beta-blocker subtypes and risk of low birth weight in newborns. J Clin Hypertens Greenwich Conn. 2018 Nov;20(11):1603–9.

2.	Tanaka K, Tanaka H, Kamiya C, Katsuragi S, Sawada M, Tsuritani M, et al. Beta-Blockers and Fetal Growth Restriction in Pregnant Women With Cardiovascular Disease. Circ J Off J Jpn Circ Soc. 2016 Sep 23;80(10):2221–6.

3.	Davis RL, Eastman D, McPhillips H, Raebel MA, Andrade SE, Smith D, et al. Risks of congenital malformations and perinatal events among infants exposed to calcium channel and beta-blockers during pregnancy. Pharmacoepidemiol Drug Saf. 2011 Feb;20(2):138–45.

4.	Cruickshank DJ, Campbell DM. Atenolol in essential hypertension during pregnancy. BMJ. 1990 Nov 10;301(6760):1103.

5.	Bullo M, Tschumi S, Bucher BS, Bianchetti MG, Simonetti GD. Pregnancy outcome following exposure to angiotensin-converting enzyme inhibitors or angiotensin receptor antagonists: a systematic review. Hypertens Dallas Tex 1979. 2012 Aug;60(2):444–50.

6.	Moretti ME, Caprara D, Drehuta I, Yeung E, Cheung S, Federico L, et al. The Fetal Safety of Angiotensin Converting Enzyme Inhibitors and Angiotensin II Receptor Blockers. Obstet Gynecol Int. 2012;2012:658310.

7.	Cooper WO, Hernandez-Diaz S, Arbogast PG, Dudley JA, Dyer S, Gideon PS, et al. Major congenital malformations after first-trimester exposure to ACE inhibitors. N Engl J Med. 2006 Jun 8;354(23):2443–51.

8.	Lennestål R, Otterblad Olausson P, Källén B. Maternal use of antihypertensive drugs in early pregnancy and delivery outcome, notably the presence of congenital heart defects in the infants. Eur J Clin Pharmacol. 2009 Jun;65(6):615–25.

9.	Rosenthal T, Oparil S. The effect of antihypertensive drugs on the fetus. J Hum Hypertens. 2002 May;16(5):293–8.

10.	Graham JW. Missing Data Analysis: Making It Work in the Real World. Annu Rev Psychol. 2009;60(1):549–76.

11.	GitHub [Internet]. [cited 2022 Mar 9]. Projects that have contributed to MoBa Genetics · folkehelseinstituttet/mobagen Wiki. Available from: https://github.com/folkehelseinstituttet/mobagen

12.	Helgeland Ø, Vaudel M, Sole-Navais P, Flatley C, Juodakis J, Bacelis J, et al. Characterization of the genetic architecture of BMI in infancy and early childhood reveals age-specific effects and implicates pathways involved in Mendelian obesity [Internet]. medRxiv; 2021 [cited 2022 Mar 9]. p. 2021.05.04.21256508. Available from: https://www.medrxiv.org/content/10.1101/2021.05.04.21256508v1

13.	Hughes A, Corfield E, Hemani G, Davies NM, Havdahl A. NIPH PsychGen and MRC IEU post-imputation QC of MoBaGenetics release 1.0, version 1, 27/10/202. :15.
